# Multi-Omics Analysis for Identifying Cell-Type-Specific Druggable Targets in Alzheimer’s Disease

**DOI:** 10.1101/2025.01.08.25320199

**Authors:** Shiwei Liu, Min Young Cho, Yen-Ning Huang, Tamina Park, Soumilee Chaudhuri, Thea Jacobson Rosewood, Paula J Bice, Dongjun Chung, David A. Bennett, Nilüfer Ertekin-Taner, Andrew J Saykin, Kwangsik Nho

## Abstract

**Background:** Analyzing disease-linked genetic variants via expression quantitative trait loci (eQTLs) is important for identifying potential disease-causing genes. Previous research prioritized genes by integrating Genome-Wide Association Study (GWAS) results with tissue-level eQTLs. Recent studies have explored brain cell type-specific eQTLs, but they lack a systematic analysis across various Alzheimer’s disease (AD) GWAS datasets, nor did they compare effects between tissue and cell type levels or across different cell type-specific eQTL datasets. In this study, we integrated brain cell type-specific eQTL datasets with AD GWAS datasets to identify potential causal genes at the cell type level.

**Methods:** To prioritize disease-causing genes, we used Summary Data-Based Mendelian Randomization (SMR) and Bayesian Colocalization (COLOC) to integrate AD GWAS summary statistics with cell-type-specific eQTLs. Combining data from five AD GWAS, three single-cell eQTL datasets, and one bulk tissue eQTL meta-analysis, we identified and confirmed both novel and known candidate causal genes. We investigated gene regulation through enhancer activity using H3K27ac and ATAC-seq data, performed protein-protein interaction and pathway enrichment analyses, and conducted a drug/compound enrichment analysis with the Drug Signatures Database (DSigDB) to support drug repurposing for AD.

**Results:** We identified 27 candidate causal genes for AD using cell type-specific eQTL datasets, with the highest numbers in microglia, followed by excitatory neurons, astrocytes, inhibitory neurons, oligodendrocytes, and oligodendrocyte precursor cells (OPCs). *PABPC1* emerged as a novel astrocyte-specific gene. Our analysis revealed protein-protein interaction (PPI) networks for these causal genes in microglia and astrocytes. We found the “regulation of aspartic-type peptidase activity” pathway being the most enriched among all the causal genes. AD-risk variants associated with candidate causal gene *PABPC1* is located near or within enhancers only active in astrocytes. We classified the genes into three drug tiers and identified druggable interactions, with imatinib mesylate emerging as a key candidate. A drug-target gene network was created to explore potential drug targets for AD.

**Conclusions:** We systematically prioritized AD candidate causal genes based on cell type- specific molecular evidence. The integrative approach enhances our understanding of molecular mechanisms of AD-related genetic variants and facilitates the interpretation of AD GWAS results.

## Background

Alzheimer’s Disease (AD) is a multifaceted neurodegenerative disorder characterized by progressive cognitive decline and memory loss[1]. AD is broadly categorized into early-onset and late-onset forms, with late-onset AD (LOAD) being the most common[2]. The genetic architecture of AD is complex, involving numerous deleterious variants distributed across various genes[2]. Among these, the *APOE* ε4 allele is recognized as the strongest genetic risk factor for late-onset AD[3]. Genome-Wide Association Studies (GWAS) have significantly advanced our understanding of the genetic basis of AD[4–9]. Early AD GWAS studies identified key loci like *CLU* and *CR1*[5]. The latest AD GWAS study has significantly expanded our understanding of the genetic basis of Alzheimer’s disease, identifying 83 genetic variants across 75 loci, including 42 newly discovered variants in European ancestry populations[4].

However, while GWAS studies are instrumental in identifying genetic variants associated with AD, they fail to elucidate the molecular and cellular mechanisms by which the variants contribute to the disease. Only a small fraction of these variants resides within coding regions, while a significant number of non-coding risk variants remain unexplained. To better understand the underlying mechanisms through which these risk variants act, recent studies have employed Expression Quantitative Trait Loci (eQTL) analyses[10–15] for following up study of GWAS results. The eQTL analyses can reveal how non-coding variants identified by GWAS influence the risk of AD through changes in gene expression[16]. Several public eQTL datasets derived from brain tissue have become available, including the Braineac dataset from the UK Brain Expression Consortium (UKBEC)[17], the Genotype-Tissue Expression (GTEx) consortium[18] and the MetaBrain dataset[10]. These datasets have enhanced the interpretation of GWAS findings by elucidating how risk variants regulate gene expression on the tissue level.

Furthermore, a few recent eQTL studies have demonstrated that these non-coding variants affect gene expression in a cell-type-specific manner, underscoring the complexity of their functional impact[11, 14]. Cell type-specific eQTLs enable researchers to determine the cell types that are most influenced by genetic variants and enable the identification of key cell types and regulatory networks involved in the disease progression, thereby offering enhanced understanding of the underlying mechanisms of diseases. Moreover, previous research has shown that GWAS-identified risk variants in non-coding regions can influence phenotypic outcomes by perturbing transcriptional gene promoters and enhancers[19]. For instance, a study has shown that the AD- associated genes *BIN1* and *PICALM* are regulated through AD risk variants that overlap with microglia-specific enhancers, which interact with the active promoters of these genes[19].

Understanding whether these genetic risk variants overlap with specific regulatory elements provides deeper insights into the cell-type-specific mechanisms underlying gene expression regulation.

In this study, we systematically integrated AD GWAS summary statistics with cell type-specific eQTL data to enhance our understanding of the genetic mechanisms underlying AD. We employed Summary Data-Based Mendelian Randomization (SMR) and Bayesian colocalization (COLOC) methods to identify and prioritize potential disease-causing genes. Our analysis included five recent AD GWAS datasets and three cell type-specific eQTL datasets derived from single-cell sequencing of AD brain samples, as well as a tissue-level metabrain eQTL dataset from previous studies. We focused on prioritizing candidate causal genes for follow-up functional studies in the future. We examined their associated variants and the possible effects on enhancers in a cell type-specific manner. By comparing our results with existing studies, we identified novel cell type-specific candidate genes and used tools such as eQTpLot to visualize their colocalization. Additionally, we used differential gene expression analysis data to investigate the associations between these novel candidate causal genes and AD pathology and cognitive function. This comprehensive approach aims to improve our understanding of AD’s genetic basis at the molecular and cellular level and identify potential therapeutic targets.

## Methods

### Datasets

We utilized summary statistics from 5 latest GWAS studies on AD involving European ancestry, downloaded from the NHGRI-EBI GWAS Catalog. As shown in Additional file 1: **Table S1**, Kunkle et al. 2019 included 21,982 AD cases, and 41,944 controls from the U.S., Canada, France, Germany, Netherlands, Iceland, U.K., Greece, and other regions, totaling 63,926 samples[6]. Jansen et al. 2019 involved 24,087 AD cases, 47,793 proxy cases, and 383,378 controls, with a total of 455,258 samples from the U.S., Norway, Sweden, U.K., and other regions (Additional file 1: **Table S1)**[7]. Wightman et al. 2021 analyzed 39,918 AD cases, 46,613 proxy cases, and 676,386 controls, with a total sample size of 762,917 from Finland, Iceland, Norway, Spain, Sweden, U.K., U.S., and other regions (Additional file 1: **Table S1)**[9].

Schwartzentruber et al. 2021 included 21,982 AD cases, 53,000 proxy cases, and 419,944 controls, totaling 472,868 samples from Greece, Canada, U.S., U.K., France, and Germany (Additional file 1: **Table S1)**[8]. Bellenguez et al. 2022 provided data on 39,106 clinically diagnosed AD cases, 46,828 proxy cases, and 401,577 controls, amounting to 487,511 samples from Portugal, Switzerland, Spain, Greece, Czech Republic, Netherlands, Sweden, U.S., Belgium, Norway, Finland, Denmark, Italy, U.K., Bulgaria, France, and Germany (Additional file 1: **Table S1)**[4].

We utilized multiple cis-eQTL datasets predominantly from individuals of European ancestry, including both tissue and cell type levels datasets in brain cortex (Additional file 1: **Table S2**). The Metabrain eQTL dataset offered a tissue-level cis-eQTL dataset derived from a meta-analysis of 14 bulk RNA-seq datasets focused on the brain cortex[10] (see Additional file 1: **Table S2**). Additionally, three cell type-specific cis-eQTL datasets were obtained from single-cell sequencing data. The research conducted by Fujita et al. 2024 provided a cell type-specific eQTL dataset sourced from the dorsolateral prefrontal cortex (DLPFC) (Additional file 1: **Table S2**)[14]. Furthermore, Bryois et al. 2021 provided a cell type-specific eQTL dataset encompassing the temporal cortex, cortex, white matter, DLPFC, and prefrontal cortex (PFC)[11] (see Additional file 1: **Table S2**). Moreover, we performed eQTL analysis and generated a cell type-specific eQTL dataset utilizing the snRNA dataset from the DLPFC region as reported by the previous study from Mathys et al., 2023[20] (Additional file 1: **Table S2**).

### eQTL analysis

To conduct eQTL analysis for the Mathys et al., 2023 snRNA dataset from the ROSMAP cohort, we generated pseudobulk expression profiles. We focused on seven main cell types (Excitatory neurons, Inhibitory neurons, Oligodendrocytes, Oligodendrocyte Progenitor Cells (OPCs), Astrocytes, Immune cells, Vasculature cells). Pseudobulk UMI count matrices for each cell type were generated by summing UMI counts per gene across all cells within each individual using Seurat (Version 5.0.1). Low-expression genes were filtered out using the ‘filterByExpr’ function from edgeR (version 3.40.2) with default parameters. The remaining pseudobulk counts were normalized using the trimmed mean of M-values (TMM) method from edgeR, and log2 counts per million (CPM) were computed and then quantile normalized with the ‘voom’ function from limma (version 3.54.2) as a previous study[14].

To identify cis-eQTLs within 1 Mb of the transcription start site of each gene, we used Matrix EQTL (version 2.3) for analysis. Bi-allelic SNPs were retained if they had a minor allele frequency >0.05, a call rate >95%, and Hardy-Weinberg equilibrium p > 10^-6 using PLINK2 as a previous study[14]. Gene expression was modeled using a linear regression with SNP allele counts and several covariates, and significance was determined by t-statistics. To account for population structure, the top 3 genotype principal components (PCs) were included as covariates as a previous study[14]. Additionally, the top 40 expression PCs, calculated within each cell type, were used to control for non-genetic structure as . Covariates including age, sex, post-mortem interval, study cohort (ROS or MAP), and total number of genes detected were also included as a previous study[14].

### Summary data-based Mendelian Randomization

We performed SMR and Heterogeneity in Dependent Instruments (HEIDI) tests to investigate pleiotropic associations between gene expression and AD within cis-regions, using the SMR software tool (version 1.3.1). The SMR method, as detailed in the previous study[21], enables the testing of whether the effect size of a SNP on a phenotype is mediated through gene expression. This tool facilitates the prioritization of candidate causal genes underlying GWAS hits for further functional studies by leveraging summary-level data from both GWAS and eQTL datasets (as mentioned above). For our analysis, we used default parameters in the SMR software with a p-value threshold of 5.0e-8 to select the top associated eQTLs for the SMR test, focusing exclusively on cis-regions. The HEIDI test, which assesses heterogeneity among dependent instruments, was conducted using a default eQTL p-value threshold of 1.57e-3 to filter SNPs for each probe, corresponding to a chi-squared value (df =1) of 10. The association between gene expression and AD was determined as P-value of SMR < 0.05/number of probes tested[21]. For the HEIDI test, significance was determined as P-value of HEIDI > 0.05 as previous studies[21].

### Bayesian colocalization analysis

We conducted colocalization analysis using the Coloc package (version 5.2.3)[22] to investigate whether AD phenotype and gene expression share common causal variants in a given region. The input data comprised SNP effect sizes and associated p-values from both the AD GWAS and eQTL datasets (as mentioned above), formatted according to the package’s requirements. Using the coloc.abf function in the package, we tested the hypothesis of a shared causal variant under the assumption of at most one causal variant per trait. Colocalization analysis calculates posterior probabilities (PPs) of the five hypotheses: 1) PPH0; no association with either gene expression or phenotype; 2) PPH1; association with gene expression, not with the phenotype; 3) PPH2; association with the phenotype, not with gene expression; 4) PPH3; association with gene expression and phenotype by independent SNVs; and 5) PPH4; association with gene expression and phenotype by shared causal SNVs. As a large PP for H4 strongly supports shared causal variants affecting both gene expression and phenotype, we considered PPH4 > 0.75 and PPH4/PPH3 >3 as strong evidence for colocalization as previous studies[23].

### Network analysis of cell type specific candidate causal genes

For each cell type, we utilized the identified candidate causal genes as input to construct a cell type-specific protein–protein interaction (PPI) network. This network was generated using STRING (version 12.0) with a confidence score threshold of 0.4 as the minimum required interaction score and default settings for all other parameters. The resulting network was then visualized using Cytoscape (version 3.10.2). In the network, nodes represent genes, proteins, or other molecular entities, while edges illustrate the interactions between these molecules.

### Pathway enrichment of all candidate causal genes

To perform pathway enrichment analysis, we utilized the all-candidate causal genes in Metascape v3.5.20240101[24]. We conducted pathway and process enrichment analyses using various ontology sources, including KEGG Pathway, GO Biological Processes, Reactome Gene Sets, Canonical Pathways, CORUM, WikiPathways, and PANTHER Pathway. The entire genome was used as the background for enrichment calculations. Terms with a p-value < 0.01, a minimum count of 3, and an enrichment factor > 1.5 (where the enrichment factor is the ratio of observed to expected counts) were selected for further analysis. To group similar terms, we calculated kappa similarity between enriched term pairs and performed hierarchical clustering based on kappa scores. Clusters were defined with a similarity threshold > 0.3. The most statistically significant term within each cluster was identified to represent that cluster. P-values were determined using the cumulative hypergeometric distribution, and q-values were adjusted for multiple comparisons using the Benjamini-Hochberg procedure in Metascape. We showed the top 10 clusters with their representative enriched terms (one per cluster) in the results.

### eQTplot analysis for visualizing colocalization

We utilized the eQTpLot (version 0.0.0.9000) R package to visualize the colocalization between AD GWAS data and eQTL data[25]. This tool enables comprehensive visualization of gene-trait interactions by generating a series of customizable plots. Using eQTpLot, we produced visualizations that highlight the overlap between AD GWAS and eQTL signals, the correlation between their p-values, and the enrichment of eQTLs among trait-significant variants.

Additionally, the tool provided insights into the linkage disequilibrium (LD) landscape of the locus and the relationship between the directions of effect for eQTL signals and colocalizing GWAS peaks, which help us to better understand the genetic relationships between gene expression and AD.

### Cell-type-specific enhancer activity analysis

GWAS risk variants located in noncoding regions can influence phenotypic outcomes by affecting transcriptional gene promoters and enhancers[19]. Clusters of enhancers, known as super-enhancers, play a vital role in regulating cell-identity genes and are key to establishing cell-type-specific gene expression patterns[19]. In this study, we evaluated the impact of disease variants on cis-gene expression in specific cell types by evaluating whether disease variants are located within or next to regulatory elements, including enhancers and promoters. A previous study highlights that although active promoters are typically conserved across different brain cell types, active enhancers show marked cell-type specificity[19]. Thus, we focused on variant-enhancer analysis. We used a publicly available dataset, including ATAC-seq, which identifies open chromatin regions, and ChIP-seq, which marks active enhancers (H3K27ac) and promoters (H3K4me3) for each brain cell type, accessed through the UCSC genome browser session

(hg19). This dataset was generated from nuclei isolated from brain tissue resected during epilepsy treatment in 10 individuals[19]. This approach allowed us to identify which enhancers are active in specific cell types, thereby elucidating the cell-type-specific effects of disease variants on gene expression.

### Druggability analysis

To identify druggable genes, we classified the identified candidate causal genes into three tiers based on druggability confidence according to a previous study[26]. Tier 1 included genes whose protein products are targets of approved small molecule, and biotherapeutic drugs were identified using manually curated efficacy target information from release 17 of the ChEMBL database.

Tier 2 comprised proteins closely related to Tier 1 targets, identified through a BLASTP search of Ensembl peptide sequences against approved drug efficacy targets. Tier 3 encompassed proteins with more distant relationships to drug targets, identified by BLASTP with ≥25% identity over ≥75% of the sequence and E-value ≤0.001. Additionally, to prioritize alternative targets for non-druggable candidate causal genes, we utilized data from EpiGraphDB to identify directly AD related interacting genes that are indicated to be druggable with Tier1 druggability[27] based on protein-protein interaction (PPI) networks (IntAct[28], STRING[29]) and with literature or xQTL evidence for AD relevance[27].

### Potential drug/compound prediction

To identify potential pharmacological drug/compound that could modulate the expression of candidate causal genes for AD, we utilized the Drug Signatures Database (DSigDB)[30]. This resource includes 22,527 gene sets and 17,389 unique compounds linked to 19,531 genes. We accessed and downloaded the annotated drug/compound gene sets from DSigDB’s official website[31]. Using the enricher function from the R package clusterProfiler (version 4.10.1), we performed enrichment analysis to explore connections between our target genes—either druggable causal genes or tier 1 interacting genes—and potential drugs, aiming for AD drug repurposing. We set an Benjamini-Hochberg adjusted p-value threshold of <0.01 to identify drugs significantly associated with these target genes. The top 10 enriched drugs/compounds were visualized using a dot plot, and an interaction network was generated with Cytoscape (version 3.10.2) to illustrate the relationships between the target genes and the enriched drugs/compounds.

## Results

### Workflow

To identify and prioritize genes associated with AD, we integrated summary-level data from GWAS with eQTL data. As shown in **Figure 1**, we incorporated data from five recent AD GWAS datasets and three cell type-specific eQTL datasets from single-cell sequencing of AD brain samples, along with a tissue-level Metabrain eQTL dataset from previous research, as described in the **Methods**. As outlined in **Figure 1**, we first employed SMR to evaluate how SNPs associated with AD risk influence gene expression. Subsequently, we used Coloc to validate the colocalization of genetic variants within specific genomic regions. We identified 33 candidate causal genes that met our rigorous criteria (**Figure 2)**. These genes were then examined across multiple cell type-specific datasets to assess their replicability. We explored how associated variants might regulate gene expression in a cell type-specific manner, utilizing previous data on cell type-specific enhancers or promoters in brain tissue. Additionally, we compared our findings with prior studies to highlight novel candidate genes with less previous support as shown in **Figure 1**. For these novel genes, we visualized colocalization results and derived differential gene expression data from earlier studies to confirm their association with AD. Finally, we assessed the druggability of the prioritized candidate causal genes to explore potential therapeutic targets.

**Figure 1.**
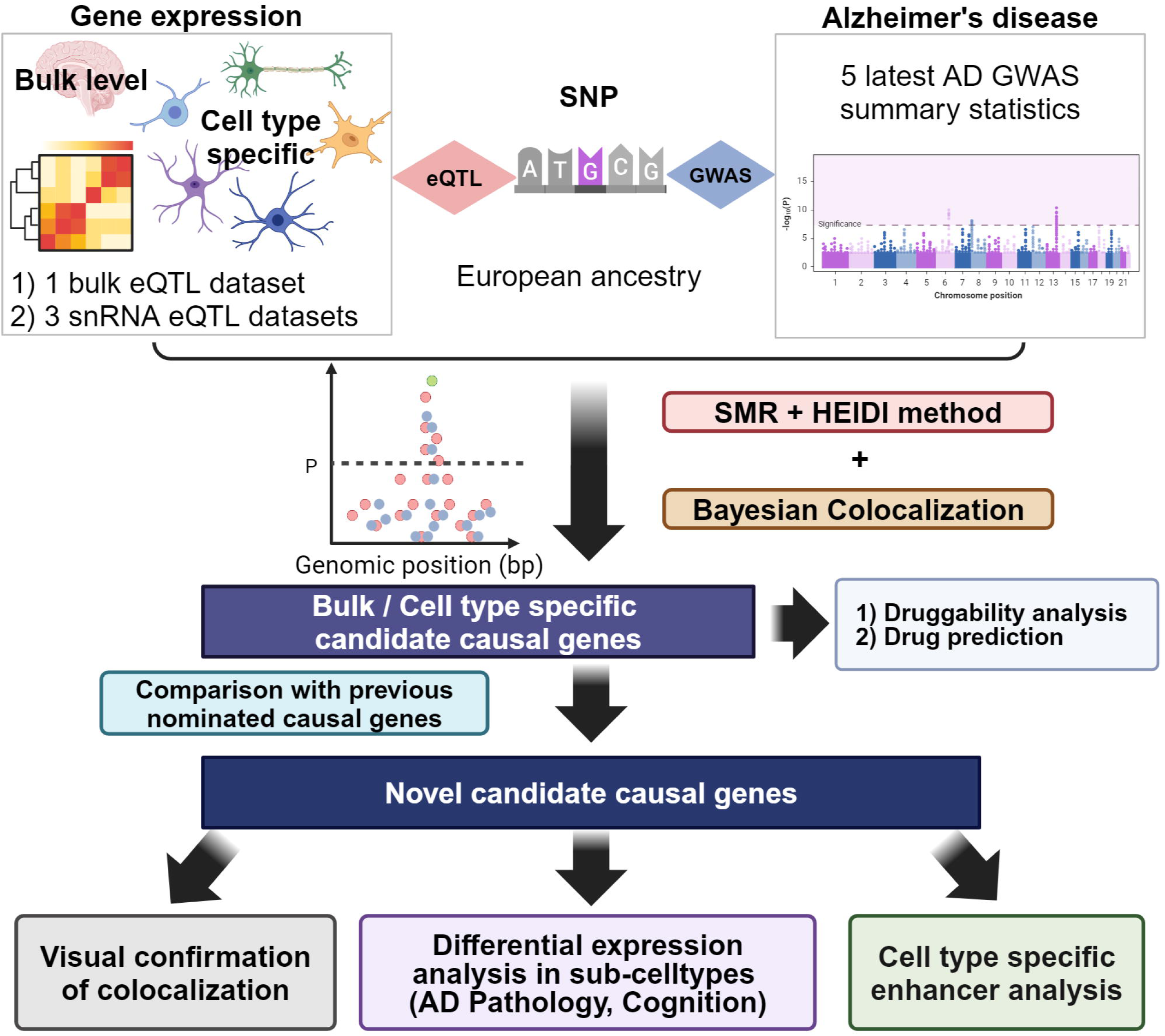
Study workflow.

**Figure 2.**
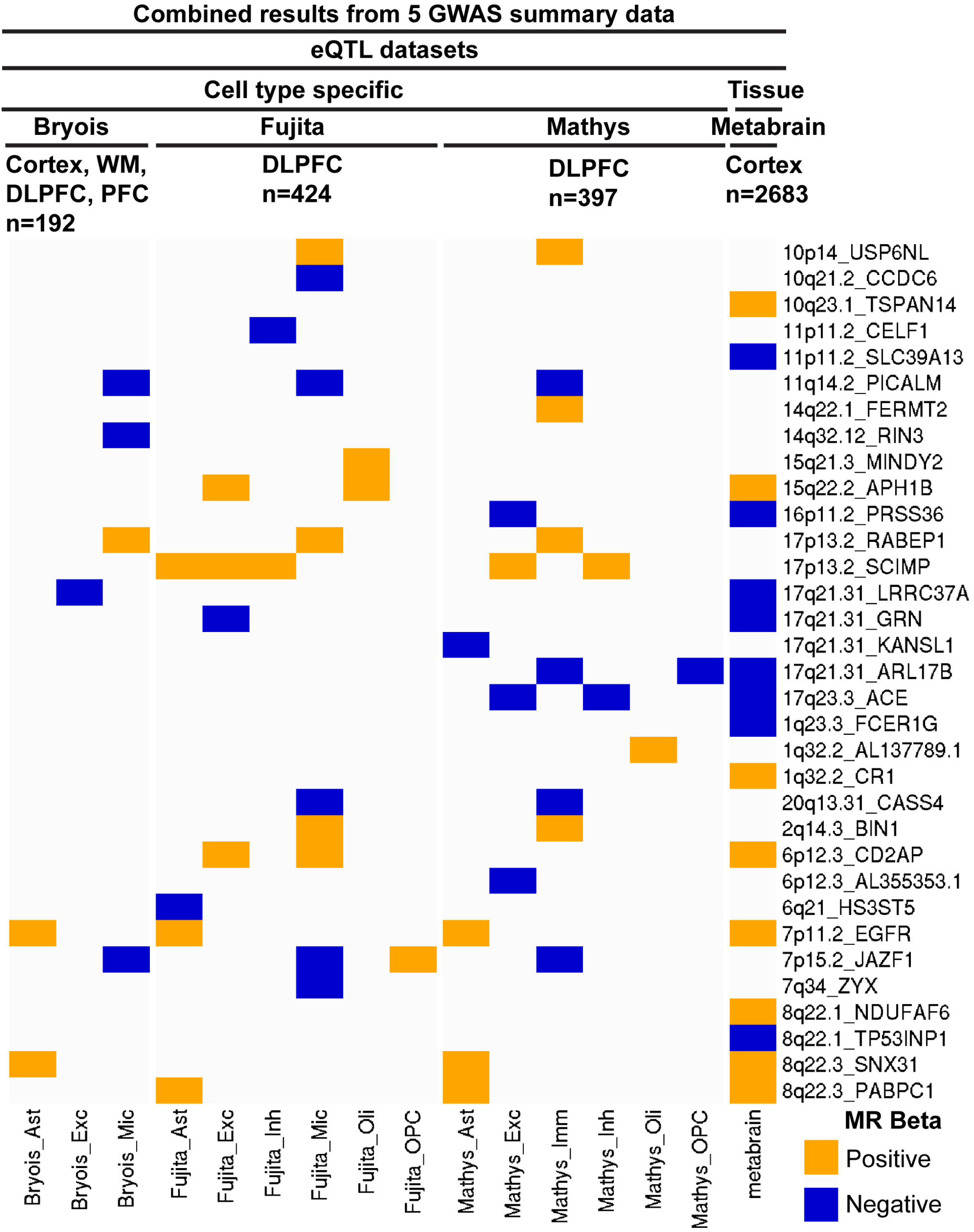
SMR beta value signs for candidate causal genes from SMR and colocalization analysis. Note: all five GWAS datasets results are combined. The candidate causal genes are filtered by SMR FDR < 0.05, HEIDI > 0.05, Coloc PPH4 < 0.75, Coloc PPH4/PPH3 > 3.

### Summary results of detected candidate causal genes

We integrated data from five recent AD GWAS datasets and three cell type-specific eQTL datasets obtained from single-cell sequencing of AD brain samples, along with a metabrain tissue-level eQTL dataset from prior research. Utilizing SMR and HEIDI as well as Coloc analyses, we identified 33 candidate causal genes across these datasets that met the filtering criteria: SMR FDR < 0.05, HEIDI p-value > 0.05, Coloc PPH4 < 0.75, and Coloc PPH4/PPH3 > 3, as shown in **Figure 2** and **Additional file 1: Table S3-S6**. Out of the 33 candidate causal genes, two (AL355353.1 and AL137789.1) are lncRNA genes, while the remaining 31 genes are mRNA genes. 27 candidate causal genes were observed in cell type-specific eQTL datasets, combining results from all GWAS datasets, as shown in **Figure 2**. As shown in **Additional file 2: Figure S1-S5**, the Bellenguez AD GWAS summary statistics revealed the highest number of candidate causal genes compared to the other AD GWAS datasets. With the combined results from all GWAS datasets, of the 27 cell type level candidate causal genes, 21 were found to be causal in only one cell type (**Figure 2)**. While genes including *ACE, CD2AP, JAZF1, APH1B*,

*ARL17B* and *SCIMP* were shared across multiple cell types, as shown in **Figure 2**. The majority consistently show the same sign in their MR beta values across different cell types. For example, *CD2AP* was detected with a positive MR beta value in both excitatory neurons and microglia in the Fujita eQTL dataset (**Figure 2)**. Interestingly, there is one gene, *JAZF1*, that exhibits an inconsistent MR beta value sign across different cell types. Specifically, *JAZF1* shows a negative MR beta value in microglia in all the Fujita, Mathys and Bryois eQTL datasets (**Figure 2)**.

However, it displays a positive MR beta value in OPCs in the Fujita eQTL dataset (**Figure 2)**. Furthermore, we noted concordant MR beta signs across single-nucleus eQTL and bulk eQTL datasets. *CD2AP*, *EGFR*, *SNX31*, *PABPC1*, *ACE*, *ARL17B*, *APH1B*, *PRSS36*, *GRN*, and *LRRC37A* are genes that are shared between the metabrain and cell type level candidate causal genes (**Figure 2)**. The MR values of these genes consistently displayed the same sign in both the metabrain dataset and the cell type level dataset (**Figure 2)**. Additionally, *TSPAN14*, *SLC39A13*, *FCER1G*, *CR1*, *NDUFAF6*, *TP53INP1* were identified exclusively as candidate causal genes in the bulk metabrain eQTL dataset (**Figure 2)**. 17 genes were identified exclusively as candidate causal genes in the cell type eQTL datasets (**Figure 2)**.

As mentioned earlier, a total of 27 candidate causal genes were observed in cell type-specific eQTL datasets (**Table 1**). The highest number of candidate causal genes was detected in microglia, followed by excitatory neurons, astrocytes, inhibitory neurons, oligodendrocytes, and OPCs (**Figure 2 and Table 1)**. We identified 10 cell type-specific candidate causal genes (*EGFR*, *SNX31*, *PABPC1*, *SCIMP*, *USP6NL*, *CASS4*, *PICALM*, *JAZF1*, *RABEP1* and *BIN1*), which were detected in at least two snRNA datasets (**Table 1**). Among these, genes *CASS4*, *PICALM*, *USP6NL*, *BIN1* and *RABEP1* were previously nominated by Agora[32, 33], while genes *JAZF1* and *SCIMP* were identified as colocalized genes in previous studies[14, 15]. *EGFR* is a recently prioritized causal gene with genetic regulation[4]. *SNX31* was identified as a colocalized gene in an earlier study with limited supporting evidence[34–36]. Additionally, *PABPC1*, located nearby *SNX31* emerged as a novel candidate causal gene with limited supporting evidence.

**Table 1.**
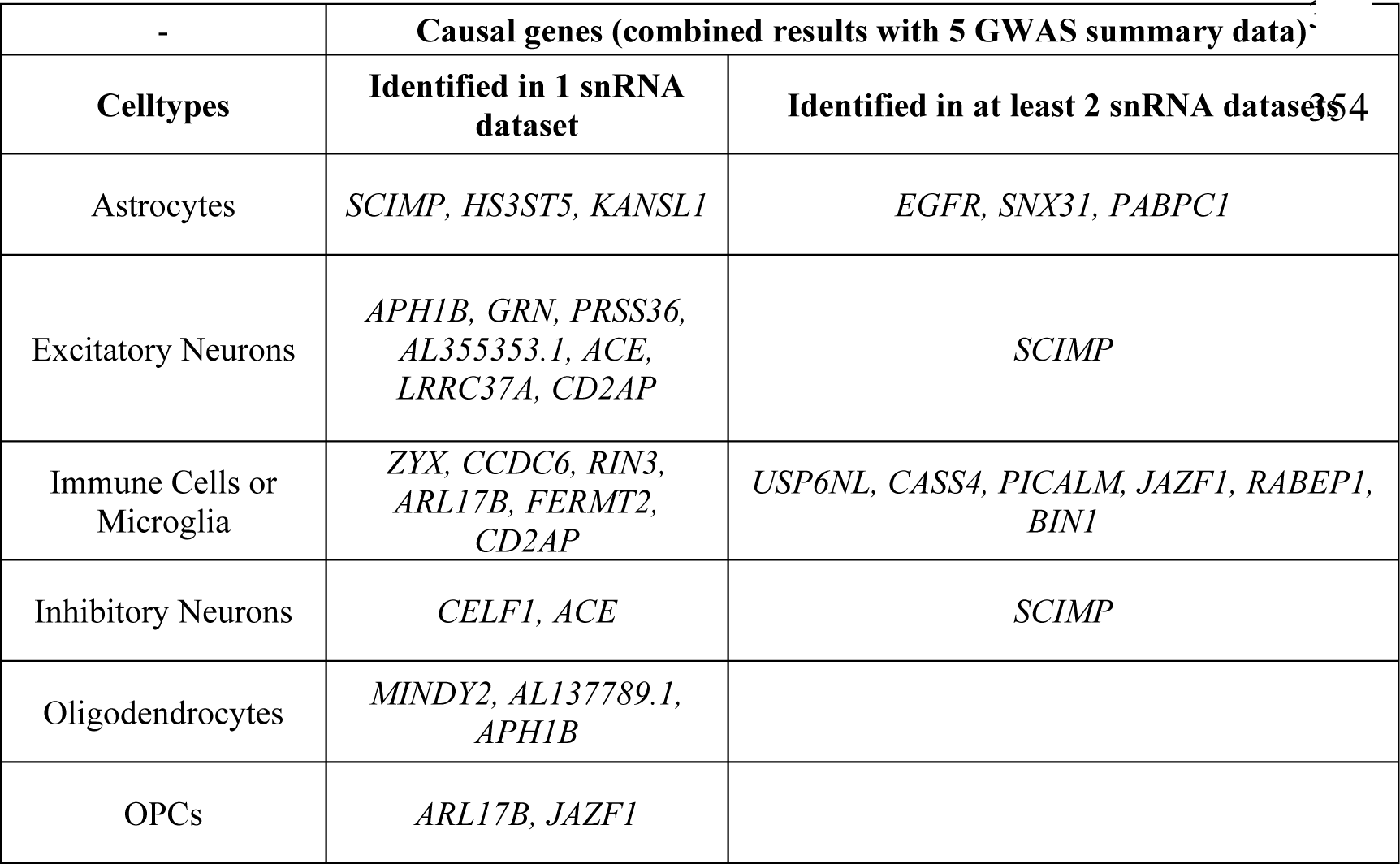
Novel discoveries, and functional analysis of candidate causal genes.

To analyze interactions among candidate causal genes for each cell type, we first constructed cell type-specific PPI networks as described in the Methods. Our PPI analysis revealed there are interactions among the corresponding proteins of the candidate causal genes in astrocytes and microglia, as illustrated in **Figure 3**. In astrocytes, we identified interactions among protein *PABPC1*, *EGFR*, and *KANSL1*, with *EGFR* serving as a central node that connects *PABPC1* and *KANSL1* (**Figure 3A)**. As shown in **Figure 3B**, the PPI network for microglia showed a more intricate interaction landscape, with 12 nodes and 16 edges. Specifically, *BIN1*, *FERMT2*, *PICALM*, *RIN3*, *CD2AP*, and *CASS4* were interconnected, indicating a complex network of interactions that could play a significant role in microglial functions related to AD.

**Figure 3.**
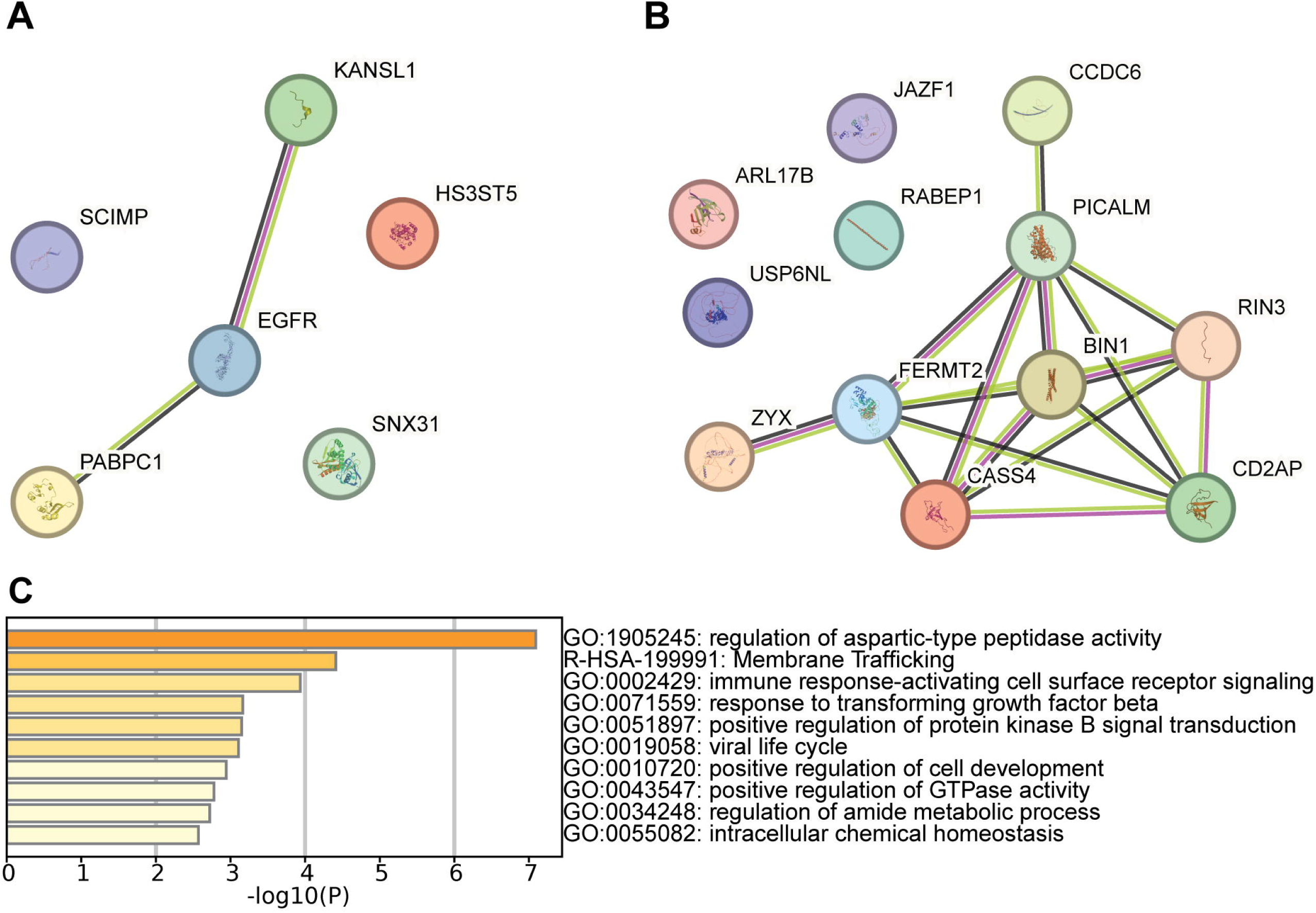
Candidate causal genes network analysis and pathway enrichment. A. STRING PPI network of Astrocyte candidate causal genes. B. STRING PPI network of Microglia candidate causal genes. C. Pathway enrichment of all 31 detected candidate causal (mRNA) genes

To identify the enriched pathways and processes, we used Metascape to perform a comprehensive enrichment analysis of the 31 candidate causal mRNA genes. **Figure 3C** displays the Top 10 clusters with their representative enriched terms, one per cluster. As displayed in **Figure 3C and Additional file 1: Table S7**, the representative enriched term of the top 1 cluster was “regulation of aspartic-type peptidase activity” (GO term) with q-value < 0.05 (q-value: 0.00178). The other clusters are not significantly enriched with q-value > 0.05.

### Visualization of colocalization for the novel astrocyte-specific candidate causal gene

We used eQTpLot to visualize the colocalization between eQTL (Astrocyte specific eQTL from Fujita et al 2024) and AD GWAS (Bellenguez et al., 2022) signals for the novel candidate causal gene, PABPC1. As shown in **Figure 4A-C**, *PABPC1* is indicated to be affected by the lead GWAS significant loci rs1693551 (GWAS P-value: 1.785e-08; Beta: 0.0459 from Bellenguez et al., 2022 AD GWAS summary statistics data). Our analysis indicates that rs1693551 may also affect the other nearby gene *SNX31* **(Figure 4B)**. We observed a tendency for eQTL to be overrepresented in the lists of significant variants from the AD GWAS (p-value = 4e-5 for *PABPC1* in astrocyte) (**Figure 4D).** Congruous SNPs effect on the gene expression in astrocyte and AD risk were also observed for *PABPC1* (**Figure 4A, 4E, 4F)**. eQTpLot P-value correlation analysis further confirms the colocalization between the *PABPC1* gene expression in astrocyte and AD risk as shown in **Figure 4E (**r = 0.81, p = 1.36e-49). The variant rs1693551 with reference allele of T and alternative allele of C is not identified as a new risk locus in the latest GWAS study[4]. However, our analysis reveals that it surpasses the genome-wide significance threshold, as illustrated by the Manhattan plot for chromosome 8 shown in **Additional file 2: Figure S6**. Additionally, we also observed colocalization of shared causal variant for *PABPC1* gene expression and AD risk with eQTL datasets from Mathys et al 2023 and Metabrain **(Additional file 2: Figure S7 and S8).** We also visualized the colocalization for the causal gene *EGFR* in astrocyte (**Additional file 2: Figure S9**).

**Figure 4.**
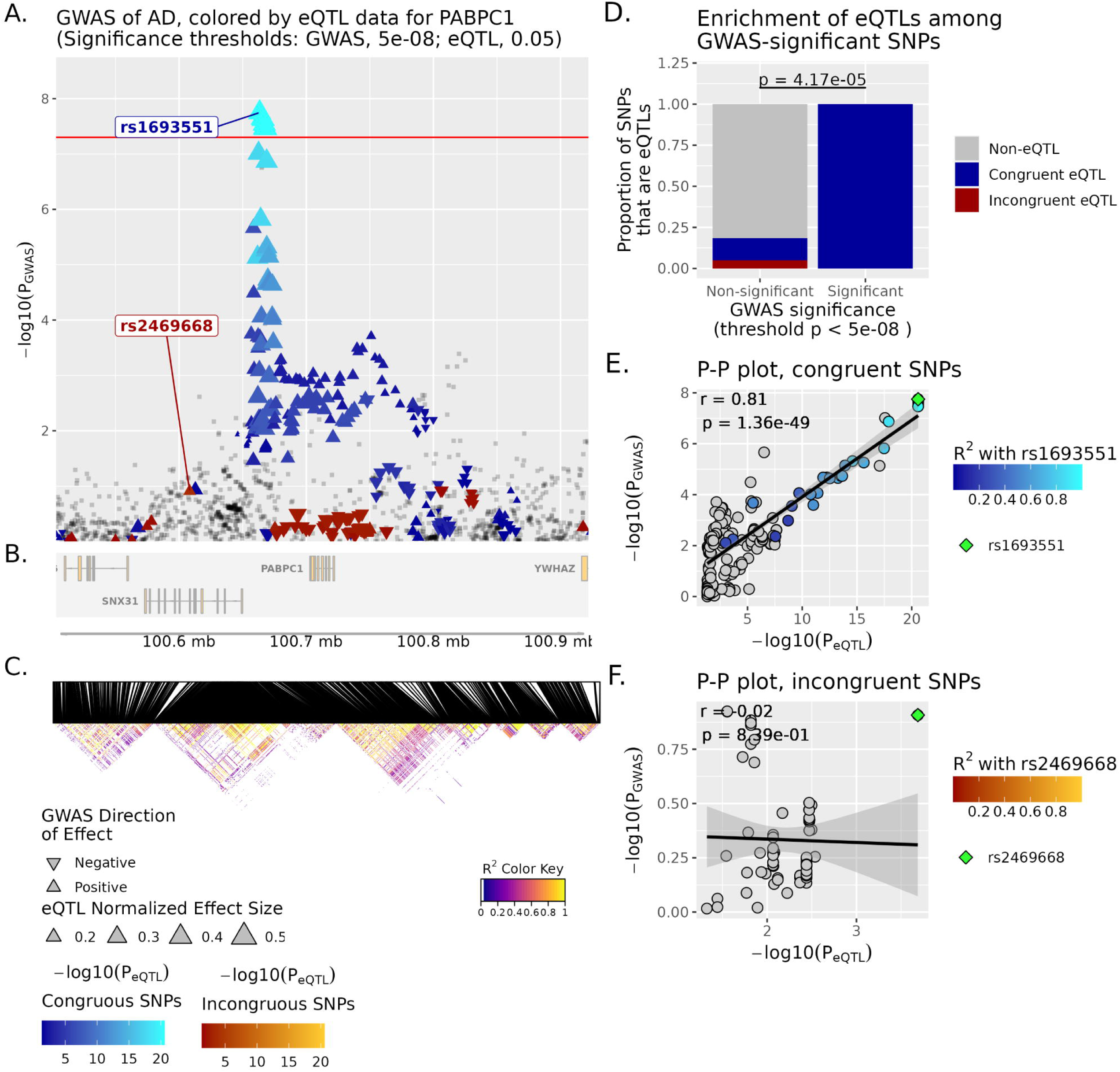
eQTpLot for colocalization between eQTLs for the gene PABPC1 and a GWAS signal for AD. The GWAS dataset is from Bellenguez et al., 2022 and the cell type eQTL dataset of astrocyte is from Fujita et al., 2024. **A** shows the locus of interest, containing the PABPC1 gene, with chromosomal space indicated along the horizontal axis. The position of each point on the vertical axis corresponds to the p-value of association for that variant with AD, while the color scale for each point corresponds to the magnitude of that variant’s p-value for association with PABPC1 expression. Variants with congruous effects are plotted using a blue color scale, while variants with incongruous effects are plotted using a red color scale. The directionality of each triangle corresponds to the GWAS direction of effect, while the size of each triangle corresponds to the effect size for the eQTL data. The default genome-wide p-value significance threshold for the GWAS analysis, 5e-8, is depicted with a horizontal red line. **B** displays the genomic positions of all genes within AD. **C** depicts a heatmap of LD information of all PABPC1 eQTL variants, displayed in the same chromosomal space as panels A and B for ease of reference (R2min=0.1, LDmin = 10). **D** depicts the enrichment of PABPC1 eQTLs among GWAS-significant variants, while **E** and **F** depicts the correlation between PGWAS and PeQTL for PABPC1 and AD, with the computed Pearson correlation coefficient (r) and p-value (p) displayed on the plot. For **E**, the analysis is confined only to variants with congruous directions of effect, while for **F** the analysis includes only variants with incongruous directions of effect. A lead variant is indicated in both **E** and **F**, and both are also labeled in panel **A**.

The MR and colocalization analyses identified a causal link between *PABPC1* gene expression in astrocytes and AD risk. To further explore this relationship, we examined *PABPC1* expression in both astrocytes and astrocyte subtypes, and its association with AD pathology and cognitive function. Specifically, we utilized differential gene expression (DEG) results from a previous study[20] focused on the DLPFC region and applied multiple testing corrections. The findings, presented in **Additional file 2: Figure S10**, indicate that *PABPC1* expression in astrocytes is significantly associated with perceptual orientation. Additionally, expression in the astrocyte sub- type GRM3 shows a suggestive association with tangle density.

### Enhancers harboring AD risk variants regulate cell-type-specific gene expression

Our results reveal that certain genes, such as *PABPC1*, was identified as candidate causal gene exclusively in astrocytes, but not in other brain cell types. This highlights that many candidate causal genes may be specific to a single cell type. To further understand this cell-type-specific effect, it is crucial to investigate how these variants influence gene expression and the underlying regulatory mechanisms. Enhancers are genomic regions that regulate gene expression, often in a cell-specific manner. A previous study [19] analyzed enhancer and promoter activity in human brain cell nuclei, revealing that genetic variants associated with brain traits and diseases exhibit cell-specific enhancer enrichment patterns. To determine if the cell-type-specific causal genes identified in our study are regulated by cell-type-specific enhancer activity, we analyzed a publicly available dataset, including ATAC-seq for open chromatin regions and ChIP-seq for active enhancers (H3K27ac) and promoters (H3K4me3) for each brain cell type, as detailed in the Methods section.

As illustrated in **Figure 5**, for the candidate causal gene *PABPC1* in astrocytes, the associated disease variant is rs1693551 (chr8, hg19_position: 10675584 bp), which is located just 59 bp from the boundary of an astrocyte-specific enhancer (chr8, hg19_position: 101675643- 101676301 bp) identified in the previous study. Given its proximity to the enhancer boundary, it is possible that the enhancer region extends beyond what was detected, especially considering the dynamic nature of enhancers and technical limitations of current detection methods. **Figure 5** shows that this enhancer, located downstream of the *PABPC1* gene, is active only in astrocytes— evidenced by prominent H3K27ac and ATAC-seq peaks—while not active in other cell types.

**Figure 5.**
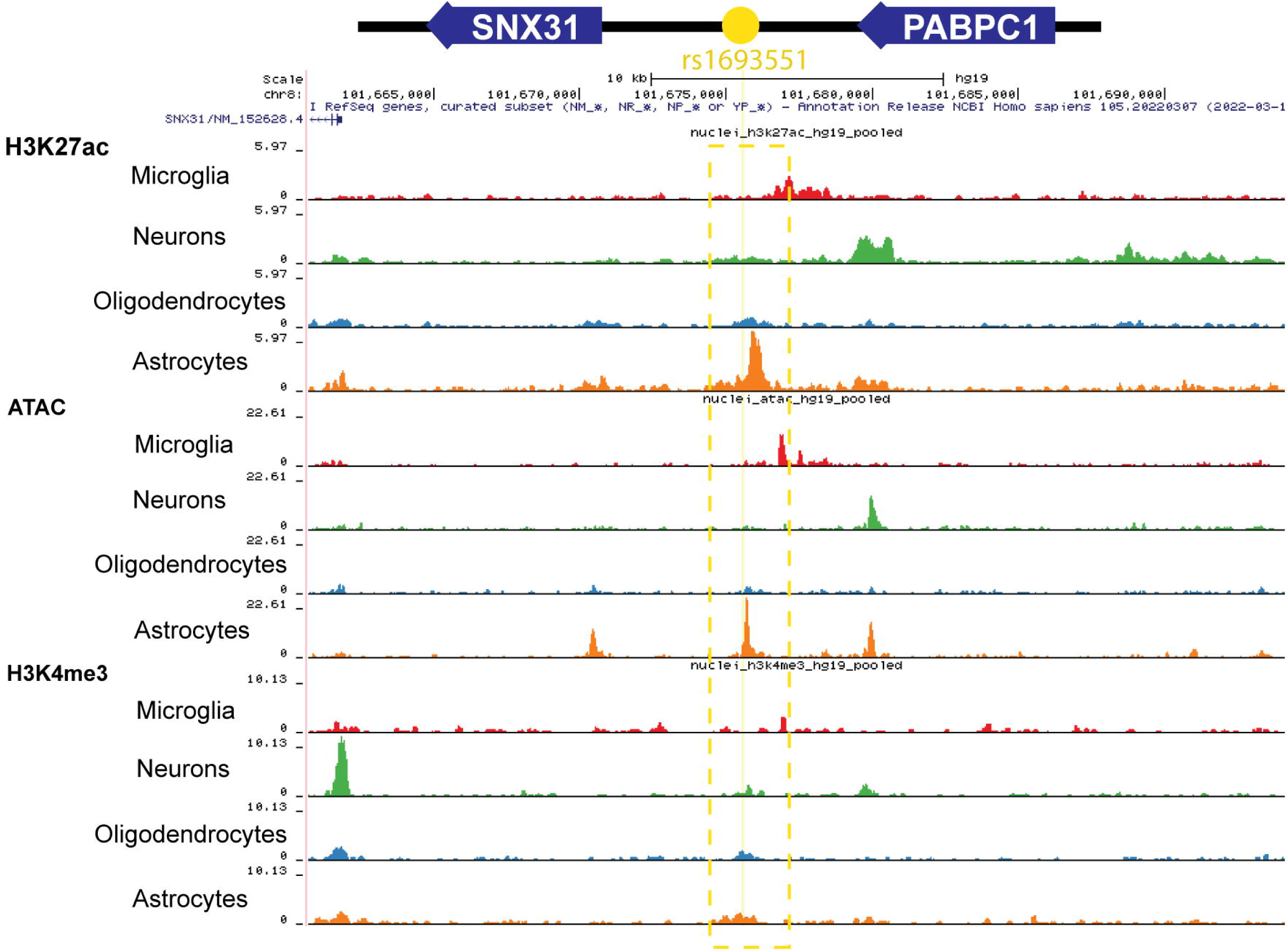
Brain cell-type-specific chromatin profiles by UCSC Genome Browser (hg19). A. H3K27ac and ATAC-seq data for *PABPC1*, showing active enhancer regions and open chromatin specific to astrocytes, with a yellow vertical line marking the location of the associated disease variant and a dashed square showing the region of active enhancer.

This suggests that the variant likely influences gene expression through a cell-type-specific enhancer, which may explain why *PABPC1* was detected as a causal gene exclusively in astrocytes.

### Druggability analysis and drug/compound prediction

To identify druggable genes from our candidate causal genes, we categorized them based on a prior drug tier classification[26]. Tier 1 includes targets of approved drugs and clinical candidates; Tier 2 includes targets with known drug-like interactions or high similarity to approved drug targets; and Tier 3 includes proteins with distant similarities to drug targets or those in key druggable families, as mentioned in the Methods. As detailed in **Additional file 1: Table S8**, we identified three candidate causal genes—*EGFR*, *ACE*, and *APH1B*—as Tier 1 druggable, and three genes—*GRN*, *PRSS36*, and *CR1*—as Tier 3 druggable. The remaining candidate causal genes were not classified as druggable based on the previous study[26]. For these non-druggable genes, we used EpiGraphDB to prioritize potential alternative drug targets within the same PPI network. We identified directly AD related interacting genes with Tier 1 druggability using PPI networks from IntAct and STRING databases, shown in **Additional file 1: Table S8**.

To identify drugs targeting the causal genes identified in this study and to broaden the scope of potential drug targets, we conducted a drug/compound enrichment analysis using DSigDB. This analysis aimed to find potential drugs for 74 target genes, which include both the druggable causal genes identified in this study and directly interacting genes with Tier 1 druggability, as detailed in **Additional file 1: Table S8**. The results of the enrichment analysis are presented in **Additional file 1: Table S9**. We focused on drugs with an adjusted p-value of less than 0.01 and selected the top 10 most significant potential drugs/compound based on their adjusted p-value (**Additional file 1: Table S9 and Figure 6A**). **Figure 6A** presents the drugs grouped by gene ratio (the percentage of target genes overlapping with the drug gene set). Within each group, the drugs are ranked by their adjusted p-value significance. The results highlight that 3-(1- methylpyrrolidin-2-yl)pyridine targets the highest number of genes, with 17 target genes including *EEF2*, *ADRB2*, *CD4*, *EGFR*, *APP*, *TFRC*, *ITGAL*, *PLD1*, *FYN*, *PIK3CA*, *RAF1*, *SRC*, *TP53*, *VEGFA*, *MAPK1*, *TNFRSF1A*, and *ACE* (**Additional file 1: Table S9**). In the second group, Imatinib mesylate is the most significant drug, targeting 14 genes, followed by Dinoprostone and Capsaicin. In the third group, histamine is the most significant drug, targeting 13 genes, followed by Gefitinib. Imatinib mesylate is detected as the most significant drug across groups. These top 10 enriched drugs (**Figure 6A**) show promise for therapeutic applications in AD and need further investigation.

**Figure 6.**
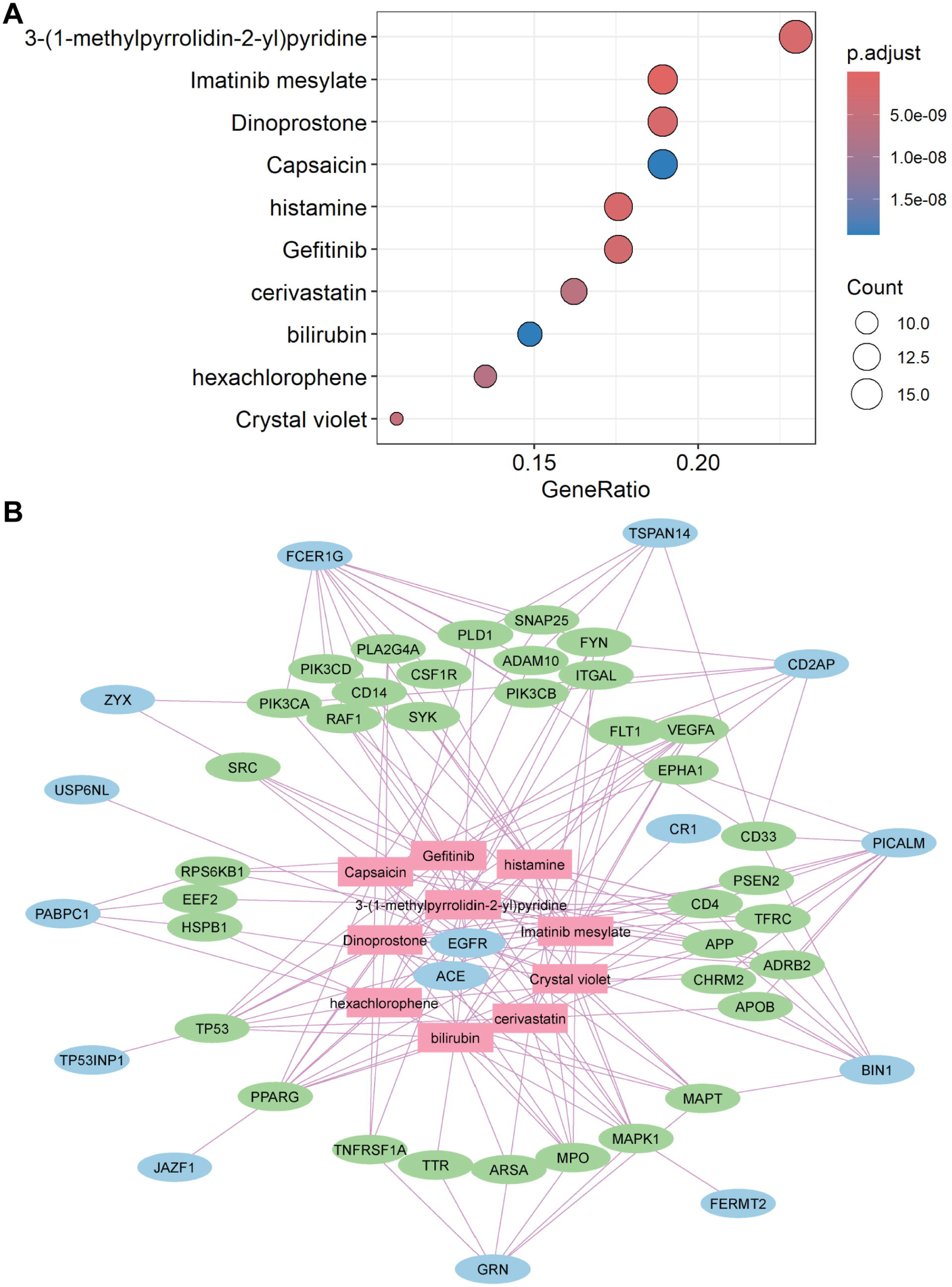
Potential drugs enrichment analysis and gene-drug interaction network. A. Top 10 enriched drug/compounds based on DSigDB predictions. B. Interaction network illustrating connections between enriched drugs/compounds and target genes. Blue circles indicate druggable/non-druggable causal genes identified in this study, green circles represent druggable interacting genes linked to non-druggable causal genes, and pink nodes denote the top 10 enriched drugs/compounds.

To illustrate the interactions between drugs and target genes—both causal genes identified in this study and directly interacting genes (AD related) with Tier 1 druggability—we constructed an interaction network using Cytoscape, as shown in **Figure 6B**. This network highlights that Tier 1 druggable genes, such as *EGFR* (targeted by all top 10 drugs) and *ACE* (targeted by 5 of the top 10 drugs) (**Additional file 1: Table S9** and **Figure 6B**), are directly targeted by multiple drugs. Additionally, the Tier 3 druggable gene *CR1* is directly targeted by Imatinib mesylate. In the network, druggable and non-druggable causal genes are represented by blue circles; interacting genes are shown in green circles, and drugs/compounds are depicted in pink (**Figure 6B**). The central area of the network features drugs and Tier 1 druggable genes, indicating direct targeting, while the surrounding groups represent interacting genes and non-druggable causal genes, which are indirectly targeted through these interactions. This visualization demonstrates the role and significance of the top 10 drugs in targeting multiple causal genes, both directly and indirectly (**Figure 6B**).

## Discussion

Many disease-associated loci exert effects that are specific to cell types[11, 14, 37, 38]. Brain diseases are influenced by genetic effects that are specific to both cell types and brain regions[11, 14, 39]. Previous GWAS studies often identify risk variants that impact disease phenotypes by regulating genes in specific tissues, yet the precise cell types involved are often not well characterized[10, 40]. Our study addresses this knowledge gap by using brain single-cell eQTL data to reveal how genetic variants impact AD at the cellular level, offering crucial insights into cell-type-specific regulation driving the disease. In this study, we combined data from five recent AD GWAS with three cell-type-specific eQTL datasets from single-cell RNA sequencing and one bulk tissue eQTL dataset from a prior meta-analysis. Through SMR and colocalization analyses, we identified candidate causal genes at both bulk and cell-type levels, uncovering novel genes and confirming known ones. We investigated gene regulation in specific cell types by analyzing enhancer activity using previous H3K27ac and ATAC-seq data. Network and pathway enrichment analyses provided additional insights into the biological relevance of these genes. To facilitate drug repurposing for AD, we performed a drug/compound enrichment analysis using the DSigDB, mapping drug interactions with both causal and interacting druggable genes. This integrated approach highlights the importance of cell-type specific functional evidence in genetic research, revealing how AD GWAS variants contribute to disease through cell-specific gene expression. By examining genetic effects at the cellular level, we gain clearer insights into AD molecular mechanism and identify promising targets for drug discovery.

In recent years, there has been growing recognition of the context-specific nature of eQTLs, extending from tissue types to functional, environmental, and cellular contexts[11, 14, 41–43]. Our study underscores the critical value of cell-type-specific eQTL datasets in identifying candidate causal genes for AD. Specifically, we identified 17 genes exclusively as candidate causal genes within the cell-type eQTL datasets (**Figure 2**). This finding highlights the limitations of bulk tissue analyses, which often aggregate signals across various cell types and may miss gene-regulatory effects that are specific to cellular contexts. By focusing on cell-type-specific eQTL data, we can uncover gene associations that are masked when only bulk tissue data is used. Furthermore, of the 27 candidate causal genes identified through cell-type-specific eQTL datasets, 21 were found to be causal in only one cell type (**Figure 2**). This cell-type specificity highlights the importance of considering cellular heterogeneity in genetic studies of complex diseases like AD.

Our study reveals that the gene JAZF1 exhibits discordant MR beta value signs across different cell types. Specifically, JAZF1 shows a negative MR beta value in microglia and a positive MR beta value in OPCs (**Figure 2**). The negative MR beta value in microglia aligns with the known downregulation of JAZF1 in multiple brain regions[44]. This discrepancy could be attributed to technical limitations, as OPCs are less prevalent in brain single-cell datasets, leading to less reliable expression measurements. However, it is also possible that the discordant MR values reflect distinct functional roles of JAZF1 in these cell types. Microglia plays a key role in immune responses and neuroinflammation, while OPCs are critical for oligodendrocyte maturation and myelination[45, 46]. The differential impact of JAZF1 on these processes could explain its varied effects across cell types. Future research should focus on validating findings in independent datasets to resolve this discordancy.

In our analysis, *PABPC1* emerged as a novel candidate causal gene for AD, highlighting its potential role in disease mechanisms. Specifically, the MR and colocalization analyses identified a causal link between *PABPC1* gene expression in astrocytes and AD risk. We found that *PABPC1* expression in astrocytes is significantly linked to perceptual orientation and shows a suggestive association with tangle density in the GRM3 astrocyte subtype. *PABPC1* is known to bind tau proteins[47]. It also regulates translation and mRNA stability[48]. Additionally, *PABPC1* is involved in stress granules and RNA splicing, critical for managing cellular stress and maintaining protein synthesis[49]. Its associations with neurofilament light chain (NF- L)[50], along with its co-localization with small tau inclusions in tauopathy[51], underscore its relevance in AD pathology. These findings warrant further investigation into *PABPC1* as a potential therapeutic target. The AD risk loci rs1693551, which achieved GWAS significance only in the latest AD GWAS summary statistics[4], has been less studied. It is the leading GWAS locus associated with the expression of the causal genes SNX31 and PABPC1 in astrocytes, underscoring its potential significance in AD. This highlights the need for further investigation into its role and relevance in the disease.

In our results, 21 of the 27-cell type level candidate causal genes were found to be causal in only one specific cell type (**Figure 2)**. Previous research indicates that cell-type-specific enhancers harboring AD-risk variants can drive such cell type-specific gene regulation[19]. For example, while *PICALM* and *BIN1* are expressed in multiple cell types, they contain microglia-specific enhancers with AD-risk variants[19]. Consistent with the previous findings, our study reveals astrocyte-specific enhancers harboring AD-risk variants associated with *PABPC1* gene expression, although interactions between these enhancers and gene promoters remain unconfirmed due to the lack of PLAC-seq data in astrocytes[19]. In addition to microglia, which are well-known for their roles in AD, our study highlights the importance of astrocytes. We provide more molecular evidence showing that astrocytes are critically involved in AD through specific gene expression and enhancer activity associated with AD-risk variants.

Our DSigDB enrichment analysis identified several drugs/compounds with potential therapeutic relevance for AD, including Imatinib mesylate, histamine, Dinoprostone, 3-(1-methylpyrrolidin- 2-yl)pyridine, Gefitinib, Crystal violet, cerivastatin, and hexachlorophene. Imatinib mesylate was highlighted as the most significant drug (**Additional file 1: Table S9**). Imatinib mesylate is notable for its role as a tyrosine kinase inhibitor and has been shown to reduce Aβ production in various experimental models[52]. Research suggests it may be effective in treating neurodegenerative disorders, including AD[53]. However, further studies are needed to fully understand its effects on the brain, particularly its ability to cross the blood-brain barrier. Some research has explored how imatinib interacts with brain transporters such as breast cancer resistance protein and P-glycoprotein[54], which is important for optimizing its use in neurodegenerative diseases. 3-(1-methylpyrrolidin-2-yl)pyridine (Nicotine) stands out for targeting the highest number of analyzed genes. Nicotine, an alkaloid in tobacco, functions by activating nicotinic acetylcholine receptors (nAChRs), which are widely expressed throughout the nervous system[55]. It has dual effects on oxidative stress and neuroprotection[56], suppresses neuroinflammation[57], and prevents Aβ aggregation[58]. Despite these benefits, its use in AD is limited by cardiovascular risks[59], addiction and negative associations with smoking[60]. However, Nicotine’s gene targeting profile found in this study suggests it could impact multiple pathways involved in AD, potentially offering a therapeutic approach through nicotinic derivatives that mitigate these adverse effects.

There are several limitations in this study. The study incorporated multiple datasets, including the three cell-type-specific eQTL datasets with partial overlap of participants from the ROSMAP cohort (see **Additional file 1: Table S2**). This partial overlap may introduce biases, potentially affecting the robustness of our findings. Furthermore, the study analyzed data from various brain regions across multiple datasets, including the cortex from the bulk metabrain eQTL dataset, the DLPFC region from the Fujita 2024 and Mathys_2023 snRNA eQTL datasets, and a range of regions such as the temporal cortex, white matter, and PFC from the Bryois 2021 snRNA eQTL dataset. The variability in brain regions might limit the generalizability of our findings, as genetic effects can be region-specific. Also, the GWAS and eQTL datasets primarily included individuals of European ancestry, which limits the generalizability of the findings to other ethnic groups. Additionally, our analysis was limited to cis-eQTLs, which reflect direct effects on genes. Cis-eQTLs do not capture the full spectrum of genetic influences, as trans-eQTLs could reveal downstream gene sets and pathways affected by disease variants. Future studies should explore available cell-specific trans-eQTL data to better understand the causal effects of genetic variants acting in trans. Furthermore, future research should use independent snRNA eQTL datasets for validation. Lastly, while our study identified potential drug targets through enrichment analysis, their clinical efficacy remains unconfirmed. Experimental validation and clinical trials are necessary to establish their therapeutic potential. Moreover, since the candidate causal genes were identified from brain tissue data and in drugs that face challenges in crossing the blood-brain barrier, further investigation is needed to evaluate the viability of these targets for drug development. In addition, despite the common challenge that smaller gene sets pose in pathway enrichment analysis due to reduced statistical power, our results with 31 input genes demonstrate that meaningful enrichments can still be detected. As shown in **Additional file 1: Table S7**, the p-value of 8.13 × 10^(-8) of the one significantly enriched pathway (regulation of aspartic-type peptidase activity) indicates a highly significant enrichment, suggesting that the observed pathway association is unlikely to have occurred by chance. Furthermore, the q-value of 1.78 × 10^(-3) of this enriched pathway confirms that the result is robust, with a very low false discovery rate, even after correcting for multiple testing. These findings indicate that, while larger gene sets generally provide more power, our analysis can still yield reliable, statistically significant results when the genes are biologically relevant.

Our analysis identified both novel and established candidate causal genes, elucidating their roles in AD molecular mechanisms and highlighting the significance of cell-type specificity in gene expression regulation and enhancer activity.

## Declarations

### Availability of data and materials

GWAS summary statistics for AD were downloaded from https://ftp.ebi.ac.uk/pub/databases/gwas/summary_statistics/GCST007001-GCST008000/GCST007511/,

https://ftp.ebi.ac.uk/pub/databases/gwas/summary_statistics/GCST013001-GCST014000/GCST013197/,

http://ftp.ebi.ac.uk/pub/databases/gwas/summary_statistics/GCST90027001-GCST90028000/GCST90027158/,

http://ftp.ebi.ac.uk/pub/databases/gwas/summary_statistics/GCST90012GCST008000/GCST007320/.

Publicly available summary statistics of metabrain eQTLs was obtained from MetaBrain website (https://www.metabrain.nl/). Fujita and Bryois Single cell eQTL datasets were obtained from Synapse: syn52335807 and https://doi.org/10.5281/zenodo.5543734, respectively. Mathys et al., 2023 snRNA dataset from ROSMAP cohort (downloaded from Synapse: syn52293442). The publicly available dataset, including ATAC-seq, which identifies open chromatin regions, and ChIP-seq, which marks active enhancers (H3K27ac) and promoters (H3K4me3) for each brain cell type, accessed through the UCSC genome browser session (hg19) at: https://genome.ucsc.edu/s/nottalexi/glassLab_BrainCellTypes_hg19

## Data Availability

All data produced in this study are available upon reasonable request to the authors. The publicly available datasets used in this study can be accessed as follows: GWAS summary statistics for Alzheimer's disease (AD) from [GCST007511](https://ftp.ebi.ac.uk/pub/databases/gwas/summary_statistics/GCST007001-GCST008000/GCST007511/), [GCST013197](https://ftp.ebi.ac.uk/pub/databases/gwas/summary_statistics/GCST013001-GCST014000/GCST013197/), [GCST90027158](http://ftp.ebi.ac.uk/pub/databases/gwas/summary_statistics/GCST90027001-GCST90028000/GCST90027158/), [GCST90012877](http://ftp.ebi.ac.uk/pub/databases/gwas/summary_statistics/GCST90012001-GCST90013000/GCST90012877/), and [GCST007320](http://ftp.ebi.ac.uk/pub/databases/gwas/summary_statistics/GCST007001-GCST008000/GCST007320/). Publicly available summary statistics of MetaBrain eQTLs are available on the [MetaBrain website](https://www.metabrain.nl/). Fujita and Bryois Single-cell eQTL datasets are available from [Synapse (syn52335807)](https://www.synapse.org/), and [Zenodo (https://doi.org/10.5281/zenodo.5543734)](https://doi.org/10.5281/zenodo.5543734). The Mathys et al. (2023) snRNA dataset from the ROSMAP cohort is available from [Synapse (syn52293442)](https://www.synapse.org/). The publicly available dataset, including ATAC-seq (identifying open chromatin regions) and ChIP-seq (marking active enhancers (H3K27ac) and promoters (H3K4me3)) for each brain cell type, can be accessed through the UCSC Genome Browser session (hg19) at [the link](https://genome.ucsc.edu/s/nottalexi/glassLab_BrainCellTypes_hg19).

## Acknowledgements

We thank the participants of the ROS/MAP study for their valuable contributions and generous donation of brain samples. We also appreciate Dr. Masashi Fujita and Dr. Hansruedi Mathys for their assistance in accessing the single-cell datasets.

## Additional files

### Additional file 1: Supplementary Tables

Table S1. Alzheimer’s disease GWAS studies. Table S2. Brain cortex region cis-eQTL datasets. Table S3. SMR and Coloc analysis results for metabrain eQTL and AD GWAS summary statistics. Table S4. SMR and Coloc results for Bryois cell type specific eQTL and AD GWAS summary statistics. Table S5. SMR and Coloc results for Fujita cell type specific eQTL and AD GWAS summary statistics. Table S6. SMR and Coloc results for Mathys cell type specific eQTL and AD GWAS summary statistics. Table S7. Pathway enrichment of candidate causal genes. Table S8. Druggability of candidate causal genes. Table S9. Drug/compound enrichment analysis results.

### Additional file 2: Supplementary Figures

Figure S1. SMR beta value and significance for candidate causal genes from SMR and colocalization analysis. Figure S2. SMR beta value and significance for candidate causal genes from SMR and colocalization analysis. Figure S3. SMR beta value and significance for candidate causal genes from SMR and colocalization analysis. Figure S4. SMR beta value and significance for candidate causal genes from SMR and colocalization analysis. Figure S5. SMR beta value and significance for candidate causal genes from SMR and colocalization analysis. Figure S6. Manhattan plot of AD GWAS (Bellenguez et al., 2022) on chromosome 8. Figure S7. eQTpLot for colocalization between eQTLs for the gene PABPC1 and a GWAS signal for AD. Figure S8. eQTpLot for colocalization between eQTLs for the gene PABPC1 and a GWAS signal for AD. Figure S9. eQTpLot for colocalization between eQTLs for the gene EGFR and a GWAS signal for AD. Figure S10: DEGs detection of PABPC1 with pathology and cognitive function.

## List of abbreviations

AD: Alzheimer’s Disease
eQTLs: Expression Quantitative Trait Loci
GWAS: Genome-Wide Association Study
SMR: Summary Data-Based Mendelian Randomization
COLOC: Bayesian Colocalization
LOAD: Late-Onset Alzheimer’s Disease
UKBEC: The UK Brain Expression Consortium
GTEx: Genotype-Tissue Expression Consortium
DLPFC: Dorsolateral Prefrontal Cortex
PFC: Prefrontal Cortex
OPCs: Oligodendrocyte Progenitor Cells
TMM: Trimmed Mean of M-values
CPM: Counts Per Million
PCs: Principal Components
HEIDI: Heterogeneity in Dependent Instruments
PPs: Posterior Probabilities
PPI: Protein–Protein Interaction
LD: Linkage Disequilibrium
DSigDB: The Drug Signatures Database
DEG: Differential Gene Expression

