## Supplementary Figures for "Multi-Omics Analysis for Identifying Cell-Type-Specific Druggable Targets in Alzheimer’s Disease"

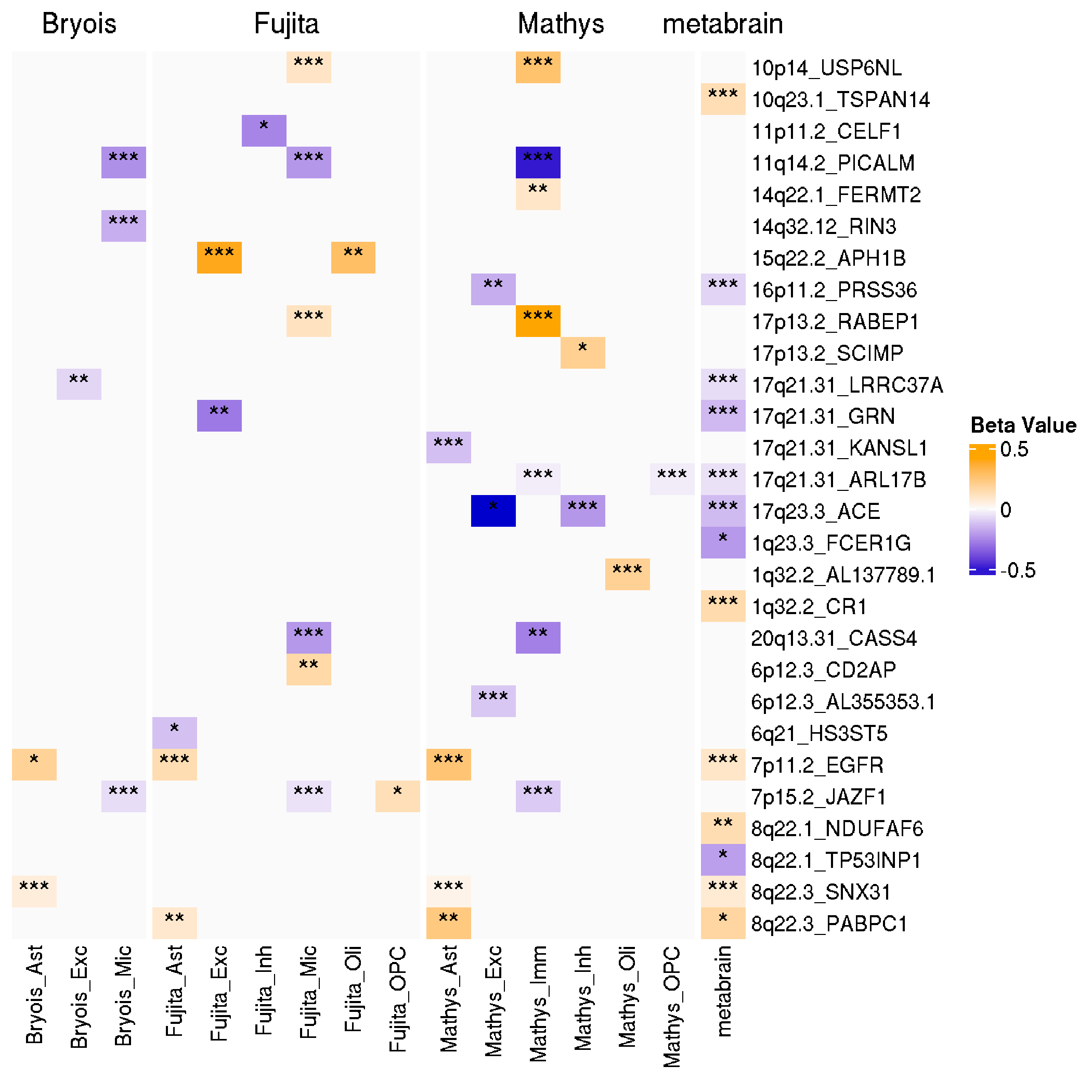

**Figure S1. SMR beta value and significance for candidate causal genes from SMR and colocalization analysis.** Note: results are based on the GWAS dataset from Bellenguez et al., 2022. The candidate causal genes are filtered by SMR FDR < 0.05, HEIDI > 0.05, Coloc PP.H4 < 0.75, Coloc PP.H4/PP.H3 > 3.

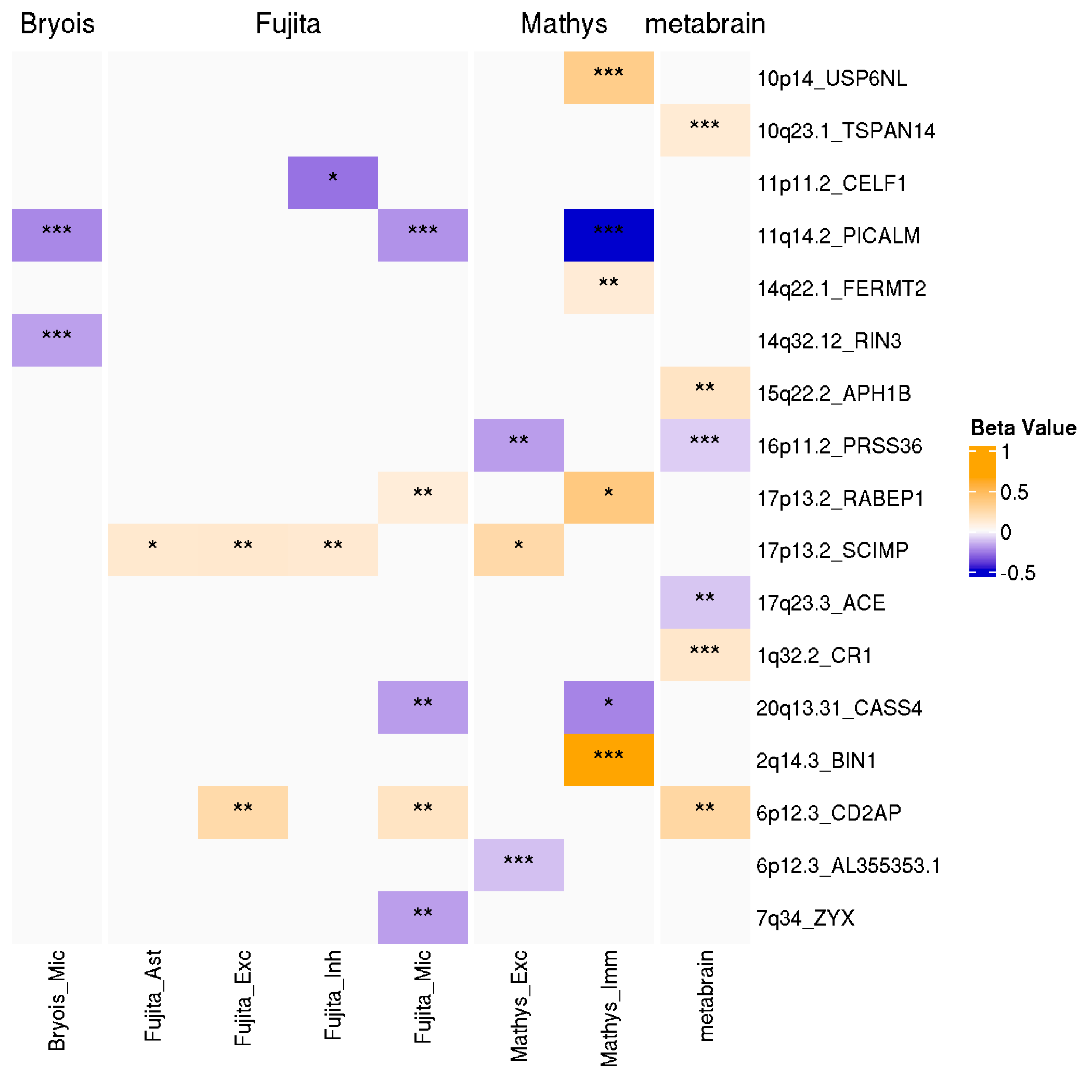

**Figure S2. SMR beta value and significance for candidate causal genes from SMR and colocalization analysis.** Note: results are based on the GWAS dataset from Schwartzentruber et al., 2021. The candidate causal genes are filtered by SMR FDR < 0.05, HEIDI > 0.05, Coloc PP.H4 < 0.75, Coloc PP.H4/PP.H3 > 3.

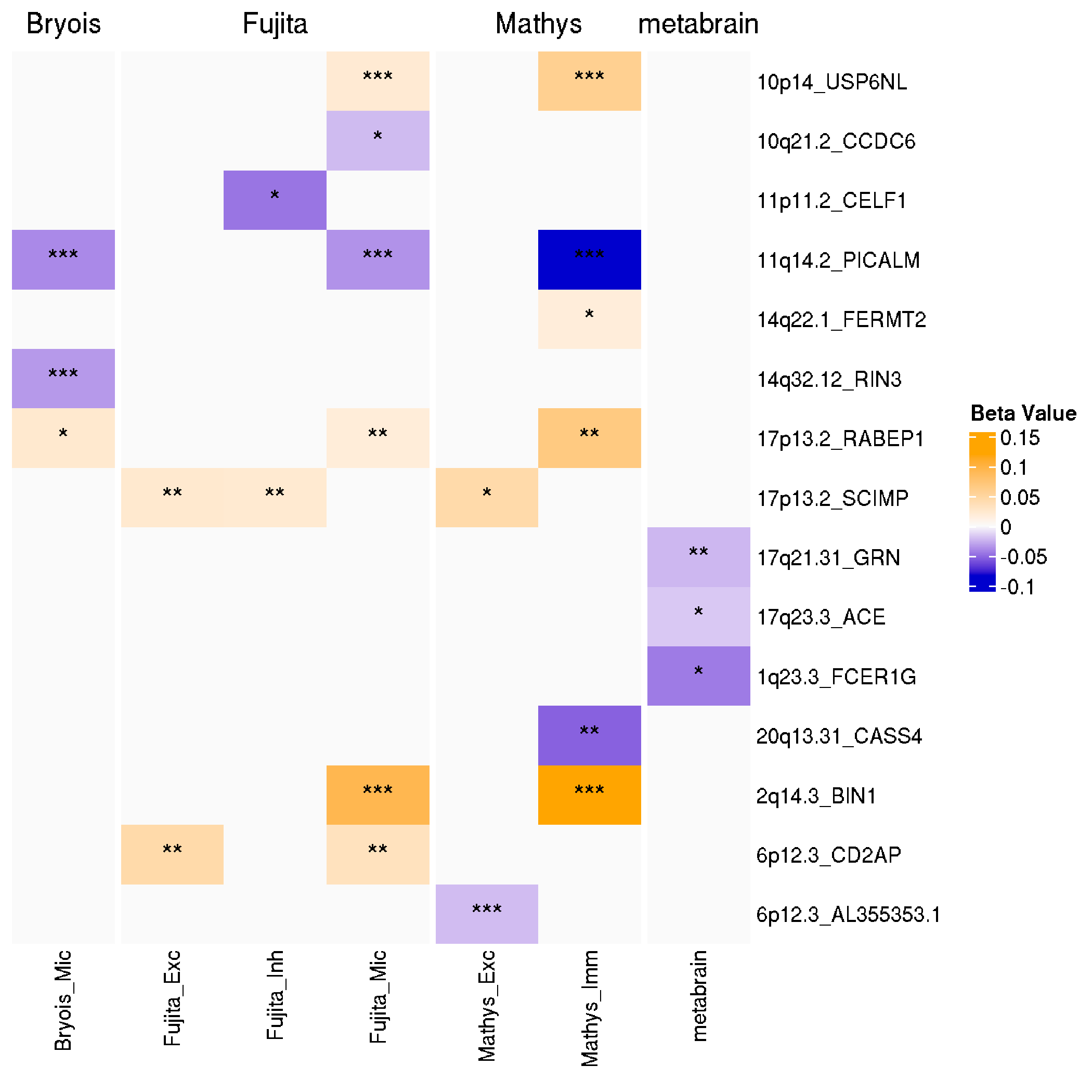

**Figure S3. SMR beta value and significance for candidate causal genes from SMR and colocalization analysis.** Note: results are based on the GWAS dataset from Wightman et al., 2021. The candidate causal genes are filtered by SMR FDR < 0.05, HEIDI > 0.05, Coloc PP.H4 < 0.75, Coloc PP.H4/PP.H3 > 3.

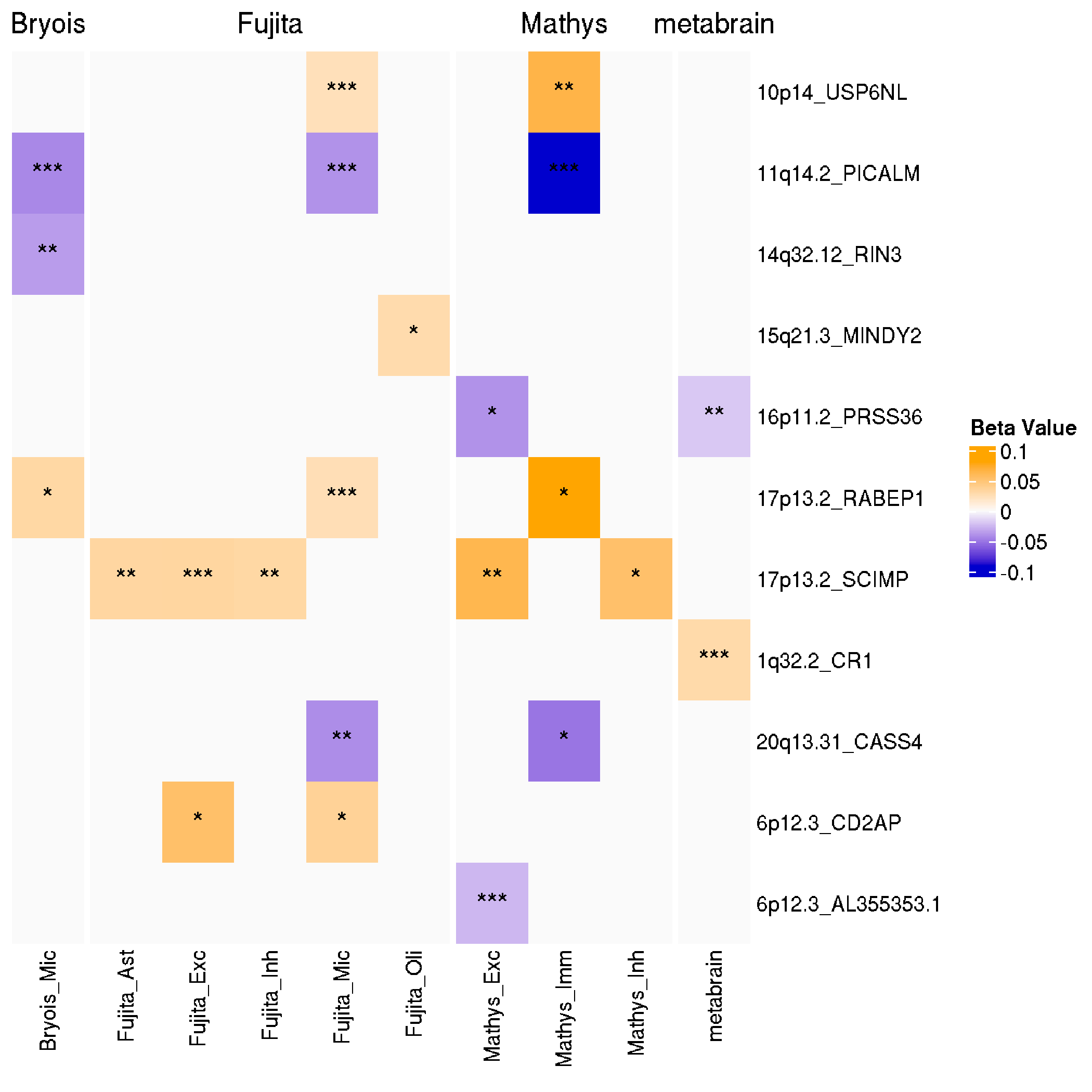

**Figure S4. SMR beta value and significance for candidate causal genes from SMR and colocalization analysis.** Note: results are based on the GWAS dataset from Jansen et al., 2019. The candidate causal genes are filtered by SMR FDR < 0.05, HEIDI > 0.05, Coloc PP.H4 < 0.75, Coloc PP.H4/PP.H3 > 3.

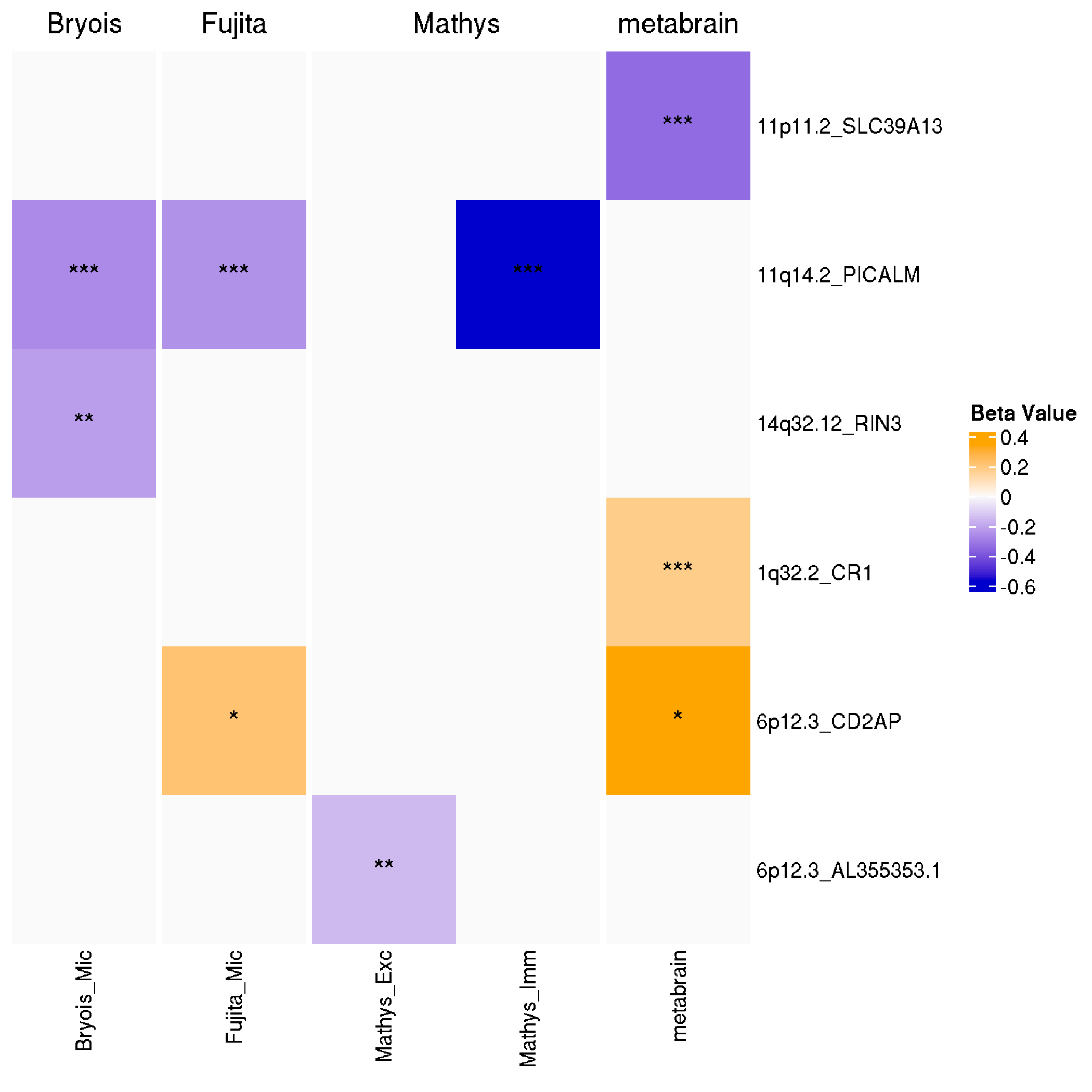

**Figure S5. SMR beta value and significance for candidate causal genes from SMR and colocalization analysis.** Note: results are based on the GWAS dataset from Kunkle et al., 2019. The candidate causal genes are filtered by SMR FDR < 0.05, HEIDI > 0.05, Coloc PP.H4 < 0.75, Coloc PP.H4/PP.H3 > 3.

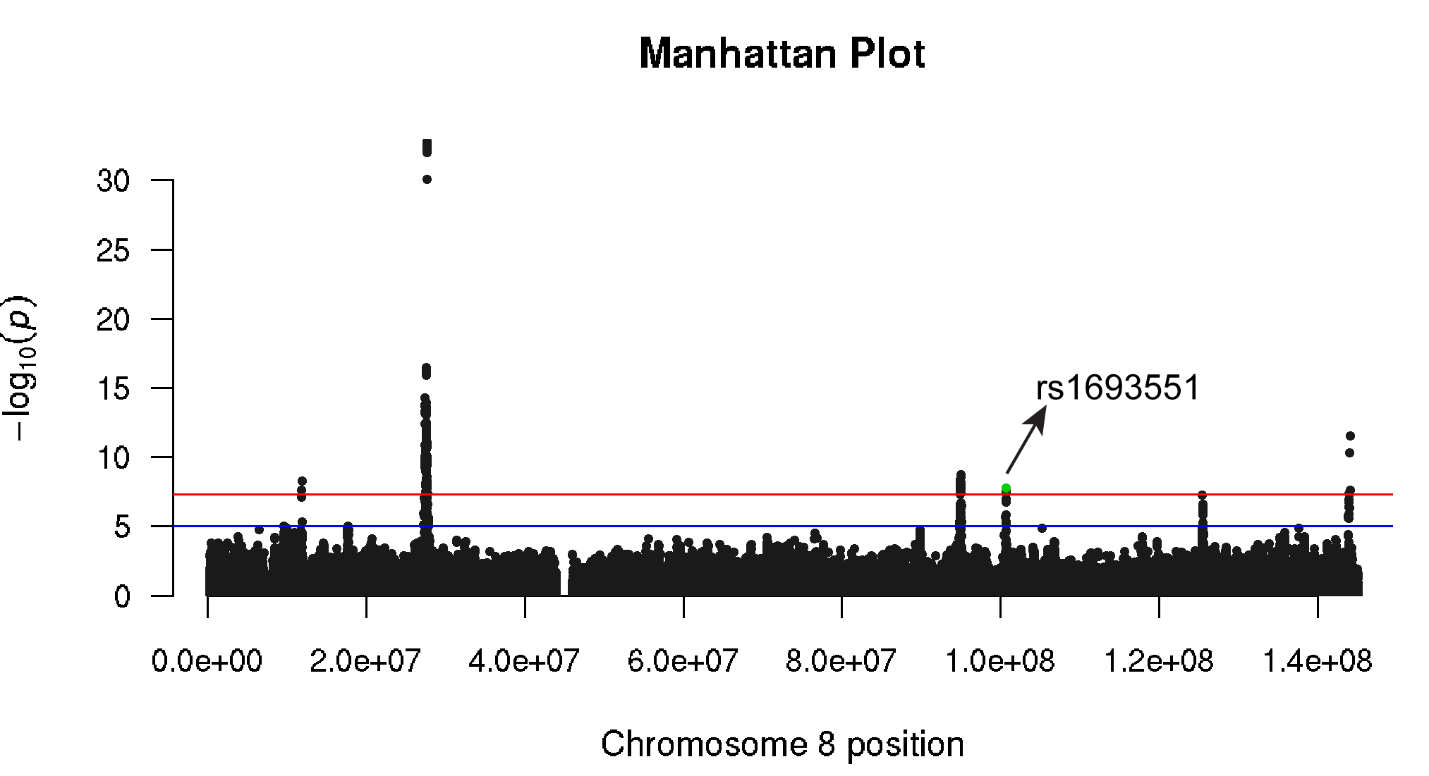
**Figure S6. Manhattan plot of AD GWAS (Bellenguez et al., 2022) on chromosome 8.**

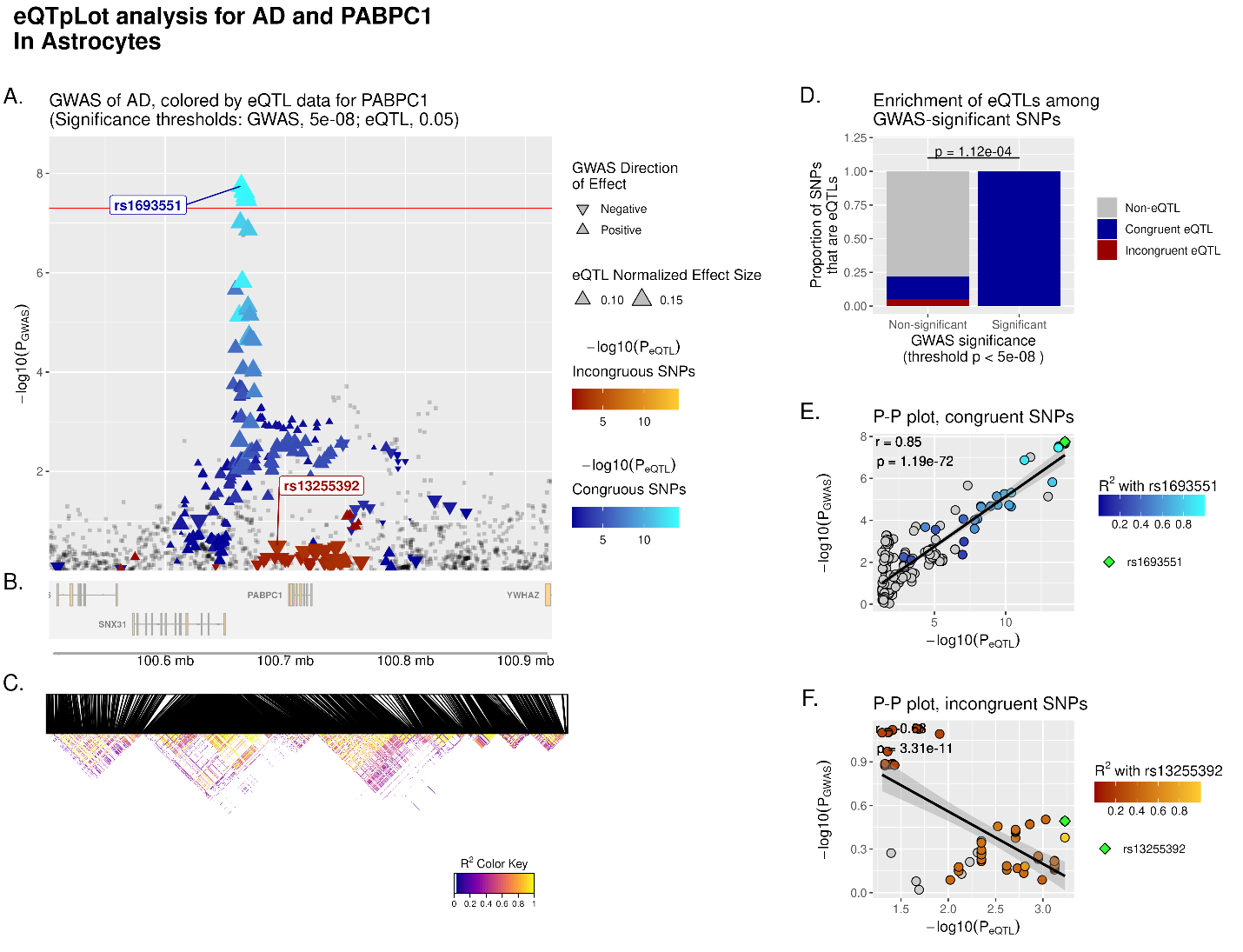
**Figure S7. eQTpLot for colocalization between eQTLs for the gene PABPC1 and a GWAS signal for AD.** The GWAS dataset is from Bellenguez et al., 2022 and the cell type eQTL dataset of astrocyte is from Mathy et al 2023. **A-F** are generated identically to **Figure 4 A-F** respectively. (R2min=0.1, LDmin = 10).

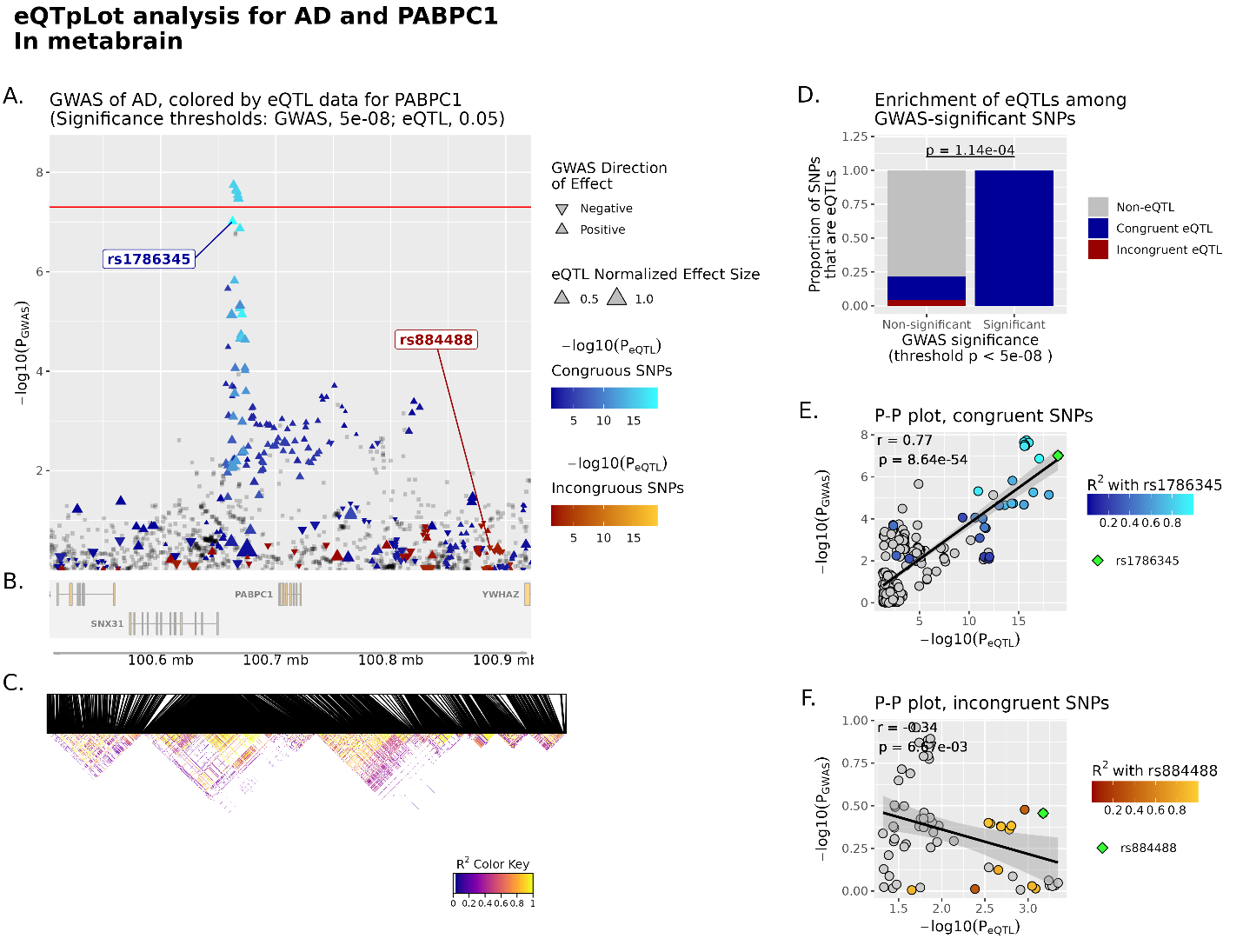
**Figure S8. eQTpLot for colocalization between eQTLs for the gene PABPC1 and a GWAS signal for AD.** The GWAS dataset is from Bellenguez et al., 2022 and the eQTL dataset is from Metabrain. **A-F** are generated identically to **Figure 4 A-F** respectively. (R2min=0.1, LDmin = 10).

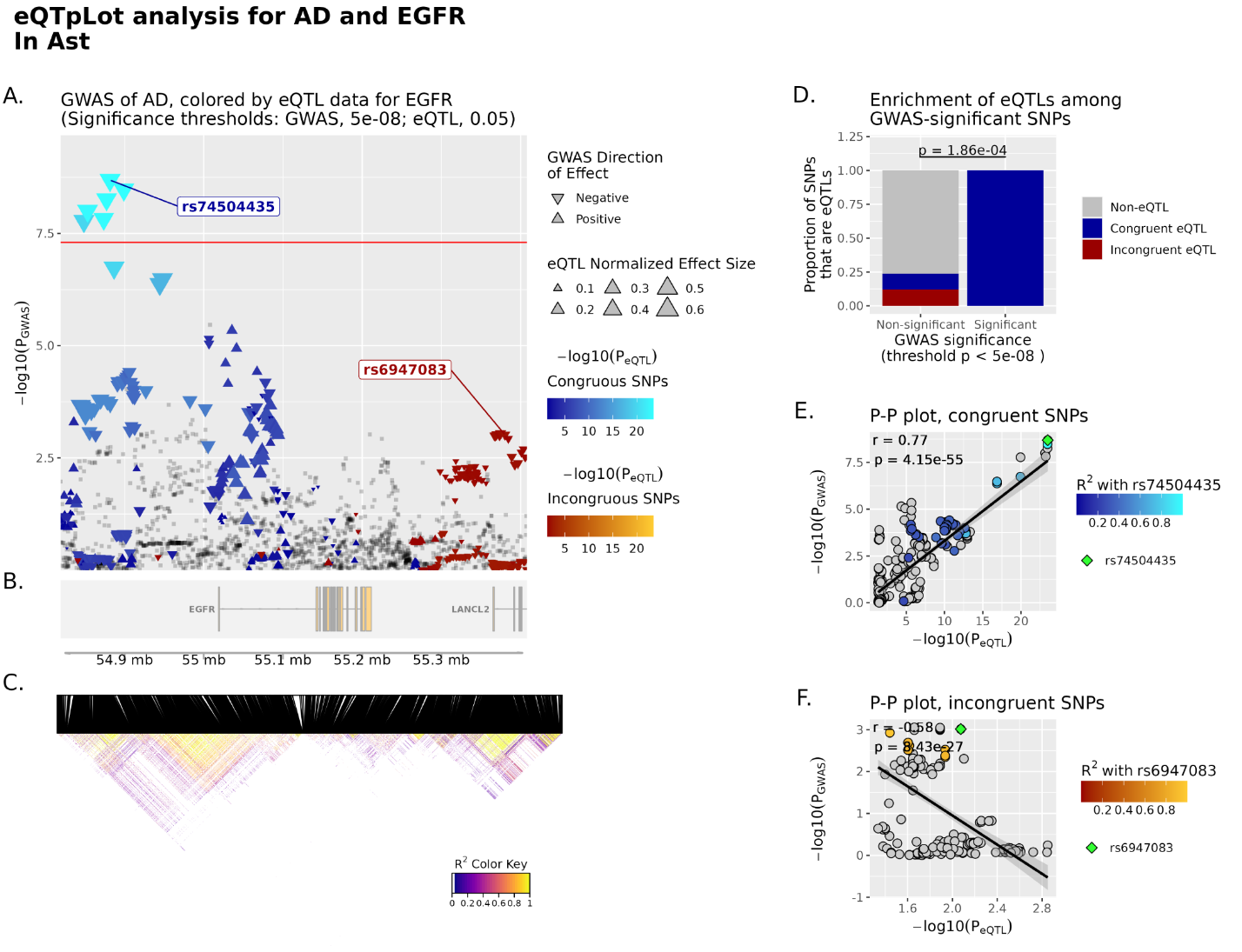

**Figure S9. eQTpLot for colocalization between eQTLs for the gene EGFR and a GWAS signal for AD.** The GWAS dataset is from Bellenguez et al., 2022 and the cell type eQTL dataset of astrocyte is from Fujita et al., 2024. **A-F** are generated identically to **Figure 4 A-F** respectively. (R2min=0.1, LDmin = 10).

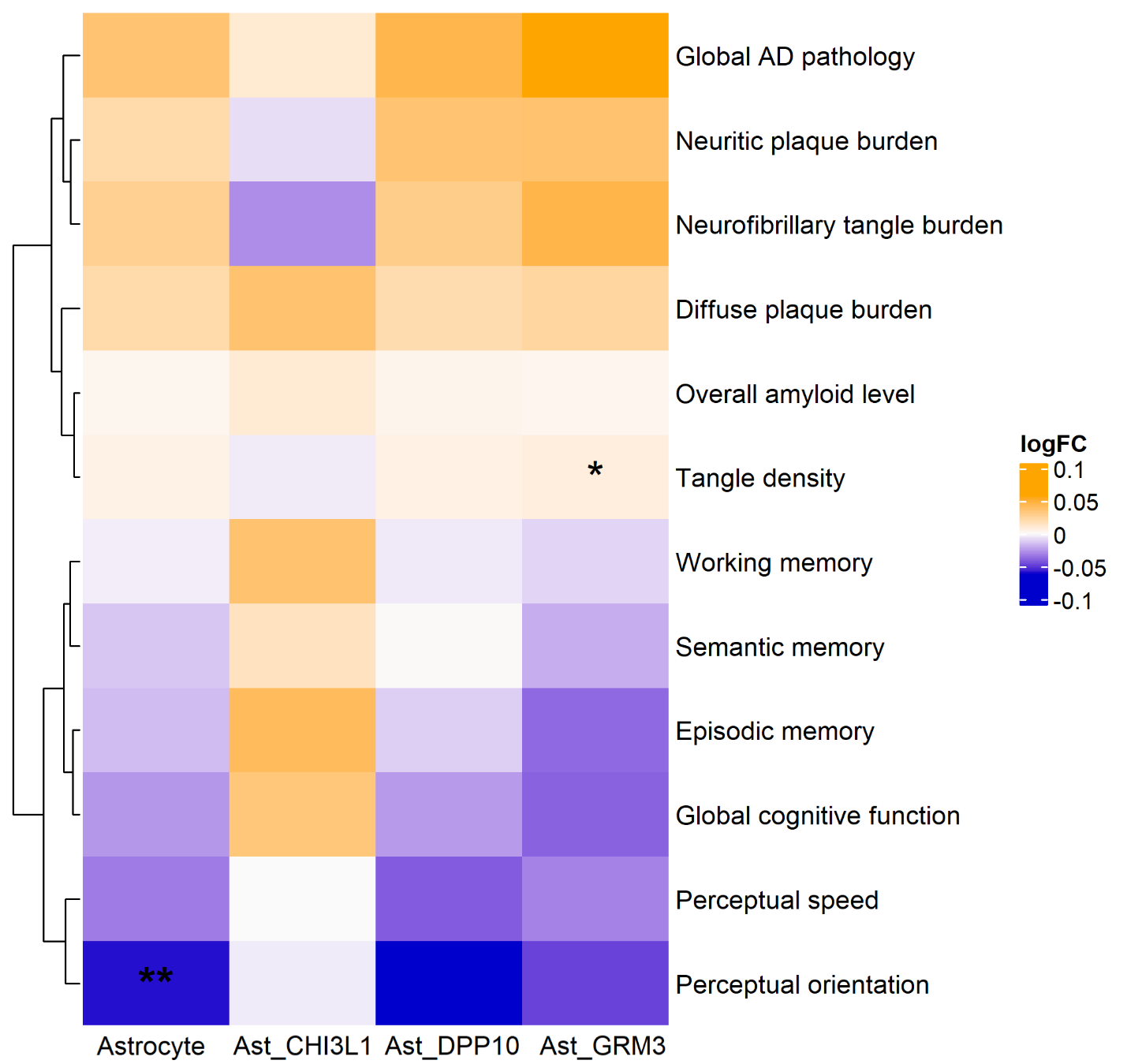
**Figure S10: DEGs detection of PABPC1 with pathology and cognitive function.**

The sign * indicates FDR < 0.1, and ** indicates FDR < 0.05
